# A Pragmatic Trial of Antibiotics and Supportive Care for Severe Pneumonia in Hospitalized Children

**DOI:** 10.64898/2026.05.05.26352430

**Authors:** Lynda Isaaka, Charles Opondo, Livingstone Mumelo, Teresiah Njoroge, Jimmy Shangala, Dennis Kimego, Rebecca Njuguna, Conrad Wanyama, Metrine Saisi, Elizabeth Isinde, Elizabeth Jowi, Achieng Adem, Julie Barasa, Mourine Ikol, Rachael Inginia, Angeline Ithondeka, Dickens Lubanga, Felicitas Makokha, Roselyn Malangachi, Christine Marete, Jecinter Modi, Maureen Muchela, Celia Wandia Kariuki, Peninnah Mwangi, Emma Namulala, Maureen Njoroge, Charles Nzioki, Sharon Ocharo, Linda Ombito, Lydia Thuranira, Magdaline Kuria, Ngina Mwangi, Esther Njiru, James Nokes, Grace Irimu, Frederick N Were, Sam Akech, Edwine Barasa, Elizabeth Maleche-Obimbo, Mike English, Elizabeth Allen, Ambrose Agweyu

## Abstract

**Background:** Evidence to guide the choice of injectable antibiotics and supportive care for children with severe pneumonia is limited and may not reflect changes in epidemiology associated with vaccination and antimicrobial resistance.

**Methods:** In this pragmatic, open-label, factorial, randomized trial conducted in 16 hospitals in Kenya, children aged 2-59 months with World Health Organization-defined severe pneumonia were assigned to receive one of three injectable antibiotic regimens: benzylpenicillin plus gentamicin (standard care), ceftriaxone, or amoxicillin-clavulanic acid. Eligible children were also randomly assigned to receive nasogastric tube feeding or intravenous fluids. The primary outcome was death from any cause by day 5 after enrollment.

**Results:** A total of 4393 children underwent randomization to the antibiotic groups, and 1064 to the supportive care groups. By day 5, deaths occurred in 87/1463 children (6.0%) receiving benzylpenicillin plus gentamicin, 82/1458 (5.6%) receiving amoxicillin-clavulanic acid (adjusted risk ratio [aRR], 0.94; 97.5% confidence interval [CI], 0.67 to 1.31), and 81/1462 (5.5%) receiving ceftriaxone (aRR vs. benzylpenicillin plus gentamicin, 0.95; 97.5% CI, 0.68 to 1.33). Death by day 5 occurred in 30/531 children (5.7%) receiving nasogastric tube feeding and 35/532 (6.7%) receiving intravenous fluids (aRR, 1.13; 97.5% CI, 0.71 to 1.79). Secondary outcomes were similar across groups.

**Conclusions:** Among children hospitalized with severe pneumonia, outcomes with benzylpenicillin plus gentamicin were similar to those with ceftriaxone or amoxicillin-clavulanic acid, and nasogastric tube feeding was similar to intravenous fluids with respect to mortality and safety.

**Trial registration:** ClinicalTrials.gov: NCT04041791; Pan African Clinical Trial Registry: PACTR202106720981298

## BACKGROUND

Pneumonia is the leading infectious cause of death in children under 5 years, responsible for more than 700,000 deaths annually, with the greatest burden in sub-Saharan Africa and South Asia (1). It is estimated that more two-thirds of childhood pneumonia cases are attributable to viral or bacterial pathogens (2). Among bacterial causes, *Streptococcus pneumoniae* and *Haemophilus influenzae type b* (Hib) are presumed to cause the most severe and fatal forms of the disease (2). Introduction and widespread uptake of the Hib and pneumococcal conjugate vaccines have reduced invasive disease caused by vaccine serotypes, (3,4). However, the incidence of pneumonia and related deaths remains high, reflecting persistent management challenges (5).

The World Health Organization (WHO) recommends treatment based on clinical severity, with severe pneumonia defined as cough or difficulty breathing plus at least one danger sign: hypoxaemia or stridor, lethargy or unconsciousness, inability to feed or persistent vomiting, or convulsions (6). First-line therapy is injectable benzylpenicillin or ampicillin plus gentamicin (7). However, changes in pathogen epidemiology following vaccine introduction and evolving patterns of antimicrobial resistance raise concerns regarding the effectiveness of standard regimens. Non-first-line antibiotics, particularly third-generation cephalosporins, are widely used in many settings despite limited evidence that they provide superior clinical outcomes. (8–10). In addition, this widespread use of broader-spectrum antibiotics has implications for antimicrobial stewardship.

Supportive care is also critical to outcomes in severe pneumonia. Severe illness is associated with increased metabolic demands and reduced oral intake resulting from respiratory distress, which may contribute to dehydration, malnutrition, and delayed recovery (11–13). There is little consensus regarding the optimal approach to hydration and nutritional support for children with severe respiratory illness, and practice varies widely. (14–17).

Intravenous fluid therapy is commonly used to provide rapid hydration but offers limited caloric content (18,19) and may increase the risk of fluid overload (20,21) or electrolyte disturbances, including hyponatraemia (22–24). In contrast, enteral feeding through a nasogastric tube provides both hydration and nutrition and may better meet the increased metabolic demands of severe illness. However, there are theoretical concerns regarding its use in critically ill children.

Severe infection and respiratory distress are associated with redistribution of blood flow away from the splanchnic circulation, which may impair gastrointestinal perfusion, reduce intestinal motility, and limit nutrient and fluid absorption. (25,26) In addition, gastric distension and delayed gastric emptying may increase the risk of aspiration.(27) These physiological considerations have contributed to uncertainty about the safety and effectiveness of enteral feeding during acute severe illness and have often led clinicians to favor intravenous fluid administration despite its lower caloric content and potential complications.

In the current trial we sought to address two clinically important questions in children hospitalized with severe pneumonia: the comparative effectiveness of commonly used injectable antibiotic regimens and the relative safety and efficacy of nasogastric tube feeding versus intravenous fluid therapy for supportive care.

## METHODS

### Trial Design and Oversight

We conducted a multicentre, factorial, open-label, pragmatic, randomized controlled trial in 16 hospitals in Kenya. Eligibility was based on bedside clinical assessment using WHO criteria, and diagnostic investigations beyond usual care were not required. Clinical management apart from the assigned trial interventions was determined by hospital teams according to national guidelines and available resources. All participating hospitals are members of the Clinical Information Network (CIN), a collaborative initiative established to improve pediatric care processes and data quality in Kenyan public hospitals (28). Trial activities were embedded within routine hospital workflows and were delivered by site clinicians working alongside the usual ward teams.

The trial protocol was approved by the Kenya Medical Research Institute Scientific and Ethics Review Unit, the University of Oxford Research Ethics Committee, and the London School of Hygiene & Tropical Medicine Ethics Committee. Regulatory authorization to conduct the trial and import study medicines was obtained from the Pharmacy and Poisons Board of Kenya. An independent trial steering committee and data and safety monitoring committee provided oversight. The first and last authors had access to all trial data and vouch for the accuracy and completeness of the data and for the fidelity of the trial to the protocol

### Participants

Children were eligible if they were 2 to 59 months of age, were admitted with WHO-defined severe pneumonia (29), and had written informed consent from a caregiver. Severe pneumonia was defined as cough or difficulty breathing accompanied by at least one danger sign, including hypoxaemia, stridor, impaired consciousness, inability to feed, persistent vomiting, or convulsions.

Exclusion criteria for the antibiotic comparison included cardiopulmonary arrest requiring resuscitation at initial presentation, contraindication or allergy to study antibiotics, referral after more than 24 hours of injectable antibiotic therapy, or prior enrolment. Additional exclusions for the supportive care comparison included absent gag reflex, inability to maintain oxygen saturation above 90% with supplemental oxygen, severe acute malnutrition, shock or severe dehydration requiring urgent resuscitation, persistent vomiting, or inability to drink or breastfeed.

### Interventions

#### Antibiotic regimens

Participants were randomly assigned to one of three injectable antibiotic regimens: benzylpenicillin (50,000 IU/kg every 6 hours) plus gentamicin (7.5 mg/kg once daily), ceftriaxone (50 mg/kg every 12 hours), or amoxicillin-clavulanic acid (30 mg/kg every 8 hours), administered intravenously or intramuscularly as appropriate. Antibiotics were administered for at least 48 hours and for up to 7 days unless discontinued or modified by the attending clinical team for clinical reasons. Children discharged before completing 7 days of injectable therapy who had improved clinically were prescribed oral amoxicillin to complete the course at home, consistent with national practice. All study antibiotics were licensed and commonly used in Kenya and were centrally procured from licensed suppliers.

#### Supportive care regimens

Eligible participants were randomly assigned to receive either intravenous fluid therapy or nasogastric tube (NGT) feeding. In the intravenous fluid group, participants received either Ringer’s lactate with 5% dextrose or 0.9% saline with 5% dextrose by continuous infusion. In the NGT-feeding group, a nasogastric tube was inserted at enrollment, and feeds were administered as a slow bolus over approximately 20 minutes every 3 hours. Acceptable feeds included breast milk, infant formula, or other age-appropriate nutrition permitted under hospital practice.

For both groups, daily maintenance volumes were calculated from admission weight using standard pediatric formulas: 100 mL/kg for the first 10 kg, 50 mL/kg for each additional kilogram up to 20 kg, and 20 mL/kg for each kilogram thereafter. The assigned supportive care intervention was continued for at least 24 hours and until the child could tolerate oral intake sufficient to meet full maintenance requirements, unless modified by the attending clinician for clinical reasons.

#### Randomisation and Allocation Concealment

Participants were randomised in a 1:1:1 ratio to the three antibiotic groups and, if eligible, in a 1:1 ratio to the two supportive care groups. The allocation sequence was computer-generated by the trial statistician at the KEMRI-Wellcome Trust Research Programme using random permuted blocks of size 4 to 6, stratified by trial site. Separate randomisation lists and sets of envelopes were prepared for the antibiotic and supportive care comparisons. Allocation was concealed using sequentially numbered, opaque, sealed envelopes. At each site, a study clinician maintained custody of the envelopes and opened them sequentially only after the participant had been enrolled.

#### Study Processes

Screening and enrolment occurred at the point of hospital admission. Depending on eligibility, participants were randomised either to the antibiotic comparison alone or to both the antibiotic and supportive care comparisons. Daily clinical assessments were performed throughout the hospital stay, with primary outcome data collected on day 5 after enrolment. Apart from the allocated study interventions, all participants received standard care as per national guidelines (30). Study clinicians and site principal investigators reviewed patients daily until discharge or death. All participants were followed up at 30 days after admission, either by telephone or, if telephone contact was not possible, through a scheduled in-person interview at the hospital.

#### Data Collection

Information collected included socio-demographic characteristics, daily clinical assessments, treatments administered, and clinical outcomes at days 5 and 30 after enrolment. Any changes to the allocated antibiotic or supportive care intervention were recorded, along with the reason for the change and any subsequent treatments prescribed. Data were initially recorded on paper case report forms (CRFs) by study clinicians and transcribed daily into a REDCap (Research Electronic Data Capture) database by study data clerks.

#### Outcomes

The primary outcome was death from any cause by day 5 after enrollment for both intervention comparisons. Secondary outcomes included death by day 30, time to full oral intake, length of hospital stay, and serious adverse events related to the interventions.

#### Sample Size

We estimated that enrolment of 4392 participants would provide 90% power to detect a 30% relative reduction in mortality (corresponding to an absolute difference of 4 percentage points) between antibiotic groups, assuming a baseline mortality of 12% (31) with a two-sided alpha level of 2.5% to account for multiple pairwise comparisons and allowing for 5% loss to follow-up. The calculation assumed no interaction between the antibiotic and supportive care interventions. The same overall sample size was expected to provide 90% power to detect a relative reduction in mortality of approximately 25% (3 percentage points) between nasogastric feeding and intravenous fluid therapy, under similar assumptions after application of eligibility exclusions for the supportive care comparison.

#### Statistical Considerations

The primary analysis followed the intention-to-treat principle. Intervention effects were estimated for comparisons of amoxicillin-clavulanic acid versus benzylpenicillin plus gentamicin, ceftriaxone versus benzylpenicillin plus gentamicin, and nasogastric feeding versus intravenous fluid therapy. Risk ratios for mortality and serious adverse events were estimated using log-binomial models, and time-to-event outcomes were analysed using Cox proportional-hazards models. Estimates were adjusted for age, trial site, and allocation to the alternate intervention where applicable.

To account for multiple comparisons in the antibiotic factor, 97.5% confidence intervals (CIs) were reported; 95% CIs were used for the supportive-care comparison. Prespecified subgroup analyses were conducted according to age, nutritional status, HIV status, malaria, and pallor. Additional sensitivity analyses, including per-protocol and complier-average causal effect analyses, are provided in the supplementary appendix.

## RESULTS

### Participants

Recruitment took place from August 22, 2019, through March 4, 2024. Enrolment was suspended between April 8, 2020, and September 1, 2021, because of the COVID-19 pandemic, with complete follow-up of enrolled participants continuing during this period.

During the recruitment period, 49,341 children were admitted to participating hospitals, of whom 5,313 met eligibility criteria. A total of 920 caregivers declined consent, and 4,393 children underwent randomization to the antibiotic comparison: 1,466 were assigned to benzylpenicillin plus gentamicin, 1,462 to amoxicillin–clavulanic acid, and 1,465 to ceftriaxone (Figure 1A). Outcome data at day 5 were available for 99.7 to 99.8% of participants across groups, and day-30 outcomes were available for 98.8 to 99.3%.

**Figure 1a.**
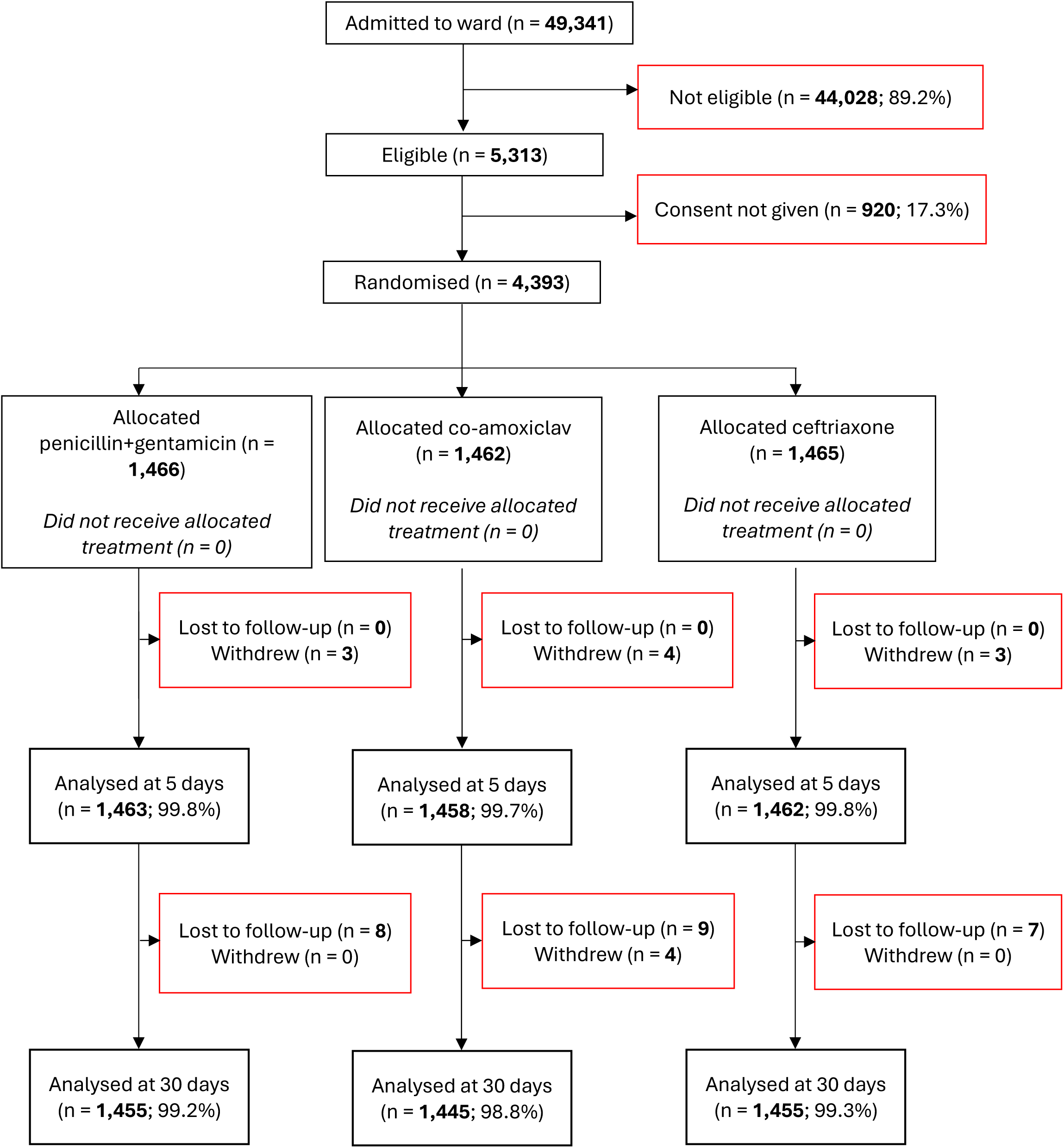
Flow diagram showing recruitment, antibiotic randomization and follow-up

Of the 5,313 children eligible for the antibiotic comparison, 1,237 also met eligibility criteria for the supportive-care comparison; 1,064 underwent randomization, with 532 assigned to nasogastric tube feeding and 532 to intravenous fluid therapy (Figure 1B). Day-5 outcome data were available for 99.8 to 100% of participants, and day-30 outcomes for 99.4% in both groups.

**Figure 1b.**
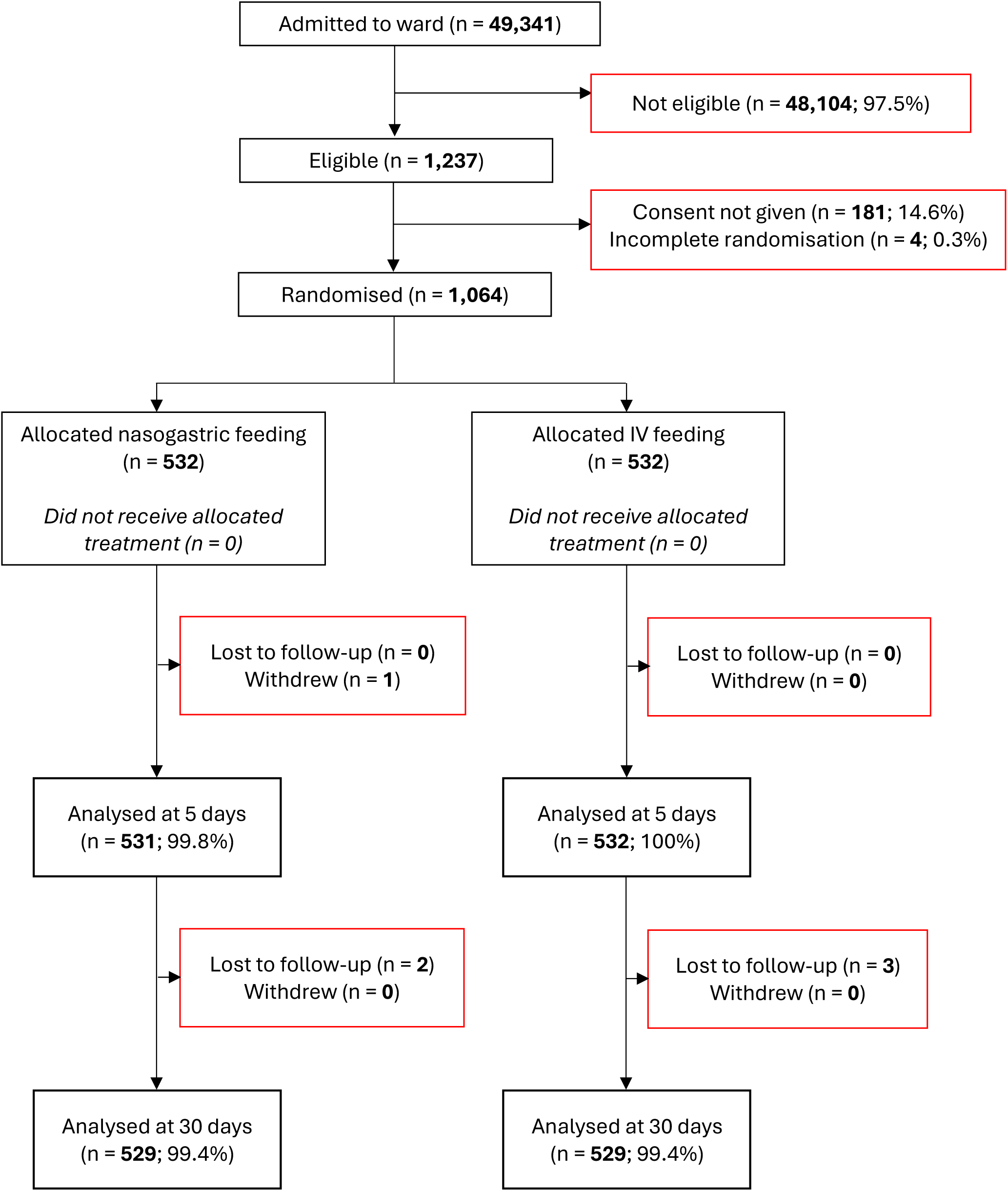
Flow diagram showing recruitment, supportive care randomization and follow-up

**Figure 2A:**
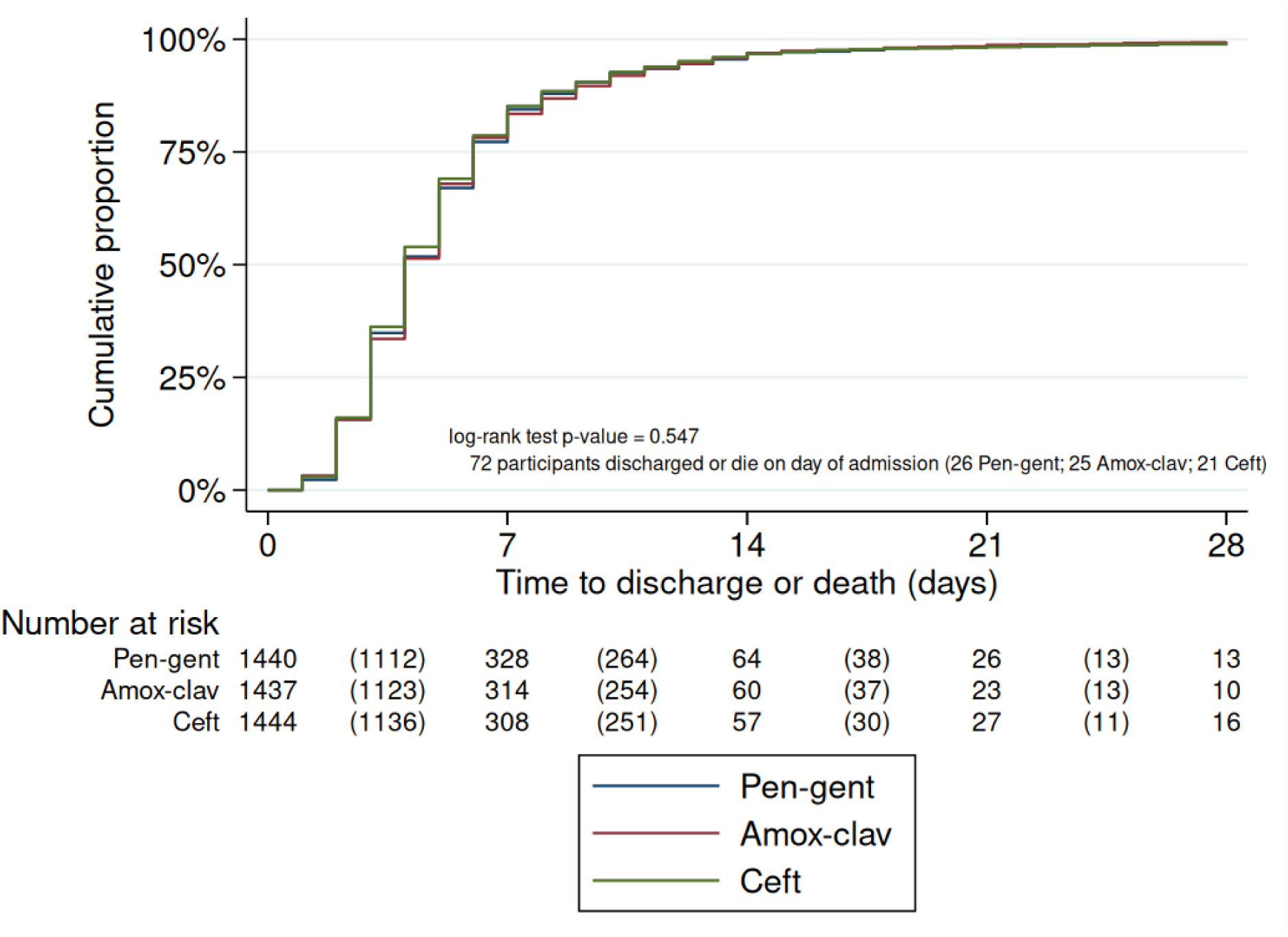
Time to discharge or death (antibiotic intervention)

**Figure 2B:**
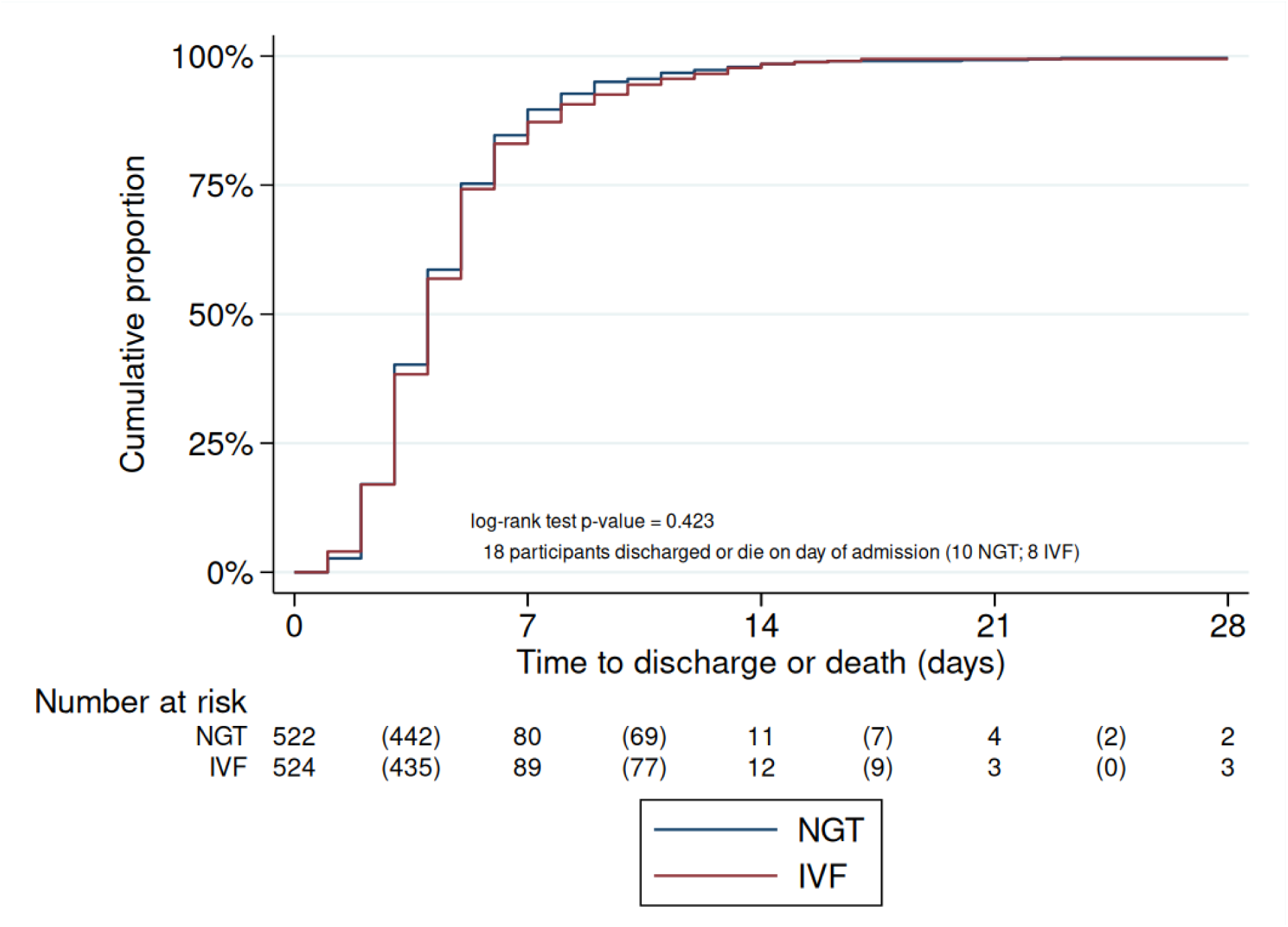
Time to discharge or death (supportive care intervention)

Baseline characteristics were similar across treatment groups (Table 1). The mean age was 14 months (SD, 12.3), 55% of participants were male, and the mean reported duration of illness before presentation was 4 days (SD, 6.5). Characteristics of eligible children who were not enrolled were similar to those of participants (Table S5 in the Supplementary Appendix).

**Table 1.** Participant characteristics by treatment arm.

|  |  | Antibiotic |  |  | Supportive Care |  |
| --- | --- | --- | --- | --- | --- | --- |
| Characteristics | Total<br>N = 4,393 | Pen-gent<br>N = 1,466 | Amox-clav<br>N = 1,462 | Ceftriaxone<br>N = 1,465 | NGT<br>N = 532 | IVF<br>N = 532 |
| **History and socio-demographic profile** |  |  |  |  |  |  |
| Sex – n (%) |  |  |  |  |  |  |
| Male | 2415 (55) | 793 (54.1) | 811 (55.5) | 811 (55.4) | 320 (60.2) | 272 (51.1) |
| Female | 1978 (45) | 673 (45.9) | 651 (44.5) | 654 (44.6) | 212 (39.8) | 260 (48.9) |
| Age 12 months or older – n (%) | 1863 (42.4) | 625 (42.6) | 593 (40.6) | 645 (44.0) | 228 (42.9) | 217 (40.8) |
| Previously hospitalised – n (%) | 441 (10) | 152 (10.4) | 139 (9.5) | 150 (10.2) | 39 (7.3) | 37 (7) |
| Discharged <1 month ago – n (%) | 66 (1.5) | 22 (1.5) | 17 (1.2) | 27 (1.8) | 6 (1.1) | 5 (0.9) |
| Length of illness (days) – mean (SD) | 4 (6.5) | 4 (4.1) | 4 (6.3) | 5 (8.4) | 4 (0.7) | 4 (0.7) |
| Diarrhoea – n (%) | 938 (21.4) | 302 (20.6) | 306 (20.9) | 330 (22.5) | 82 (15.4) | 81 (15.2) |
| Difficulty feeding – n (%) | 1928 (43.9) | 649 (44.3) | 658 (45) | 621 (42.4) | 472 (88.7) | 478 (89.8) |
| Convulsions – n (%) | 126 (2.9) | 36 (2.5) | 47 (3.2) | 43 (2.9) | 20 (3.8) | 10 (1.9) |
| Pneumococcal vaccine doses – mean (SD) | 3 (0.6) | 3 (0.6) | 3 (0.6) | 3 (0.6) | 3 (0.5) | 3 (0.5) |
| Antibiotics prior to admission – n (%) | 3628 (82.6) | 1230 (83.9) | 1192 (81.5) | 1206 (82.3) | 448 (84.2) | 439 (82.5) |
| **Physical examination** |  |  |  |  |  |  |
| Weight (kg) – mean (SD) | 8 (3.1) | 8 (3.0) | 8 (3.1) | 8 (3.2) | 9 (1.6) | 8 (1.6) |
| Height (cm) – mean (SD) | 73 (12.8) | 73 (12.8) | 72 (13) | 73 (12.7) | 73 (13.7) | 72 (13.5) |
| Wasting – n (%) | 1076 (24.5) | 353 (24.1) | 365 (25) | 358 (24.4) | 74 (13.9) | 74 (13.9) |
| Temperature (Celsius) – mean (SD) | 37.6 (1.0) | 37.6 (1.0) | 37.6 (1.0) | 37.6 (1.0) | 37.6 (7.1) | 37.6 (7.1) |
| Tachypnoea – n (%) | 973 (22.1) | 322 (22) | 324 (22.2) | 327 (22.3) | 157 (29.5) | 169 (31.8) |
| Desaturation < 90% - n (%) | 2970 (67.6) | 988 (67.4) | 1009 (69) | 973 (66.4) | 335 (63) | 356 (66.9) |
| Central Cyanosis – n (%) | 55 (1.3) | 15 (1.0) | 21 (1.4) | 19 (1.3) | 9 (1.7) | 11 (2.1) |
| Indrawing – n (%) | 4221 (96.1) | 1406 (95.9) | 1408 (96.3) | 1407 (96) | 516 (97) | 511 (96.1) |
| Grunting – n (%) | 2517 (57.3) | 806 (55) | 841 (57.5) | 870 (59.4) | 331 (62.2) | 367 (69) |
| Wheezing – n (%) | 1042 (23.7) | 357 (24.4) | 345 (23.6) | 340 (23.2) | 105 (19.7) | 127 (23.9) |
| Crackles – n (%) | 3510 (79.9) | 1163 (79.3) | 1162 (79.5) | 1185 (80.9) | 386 (72.6) | 404 (75.9) |
| Capillary refill 2 seconds or more – n (%) | 998 (22.7) | 322 (22.0) | 337 (23.1) | 339 (23.1) | 101 (19.0) | 104 (19.5) |
| Pallor – n (%) | 1098 (25.0) | 369 (25.2) | 353 (24.2) | 376 (25.7) | 125 (23.5) | 117 (22.0) |
| Can drink / breastfeed – n (%) | 2832 (64.5) | 920 (62.8) | 952 (65.1) | 960 (65.5) | 15 (2.8) | 17 (3.2) |
| Reduced level of consciousness – n (%) | 265 (6.0) | 94 (6.4) | 91 (6.3) | 80 (5.5) | 61 (11.5) | 54 (10.2) |

Pen-Gent - benzylpenicillin plus gentamicin; Amox-clav – amoxicillin clavulanic acid; NGT
– Nasogastric tube; IVF – Intravenous fluid
**Note: Percentages are column percentages (i.e. overall and within-group prevalence).**

### Primary Outcome

By day 5, deaths occurred in 87/1,463 participants (6.0%) assigned to benzylpenicillin plus gentamicin, 82/1,458 (5.6%) assigned to amoxicillin-clavulanic acid (adjusted risk ratio [aRR], 0.94; 97.5% confidence interval [CI], 0.67 to 1.31), and 81/1,462 (5.5%) assigned to ceftriaxone (aRR vs. benzylpenicillin plus gentamicin, 0.95; 97.5% CI, 0.68 to 1.33) (Table 2A).

**Table 2a.**
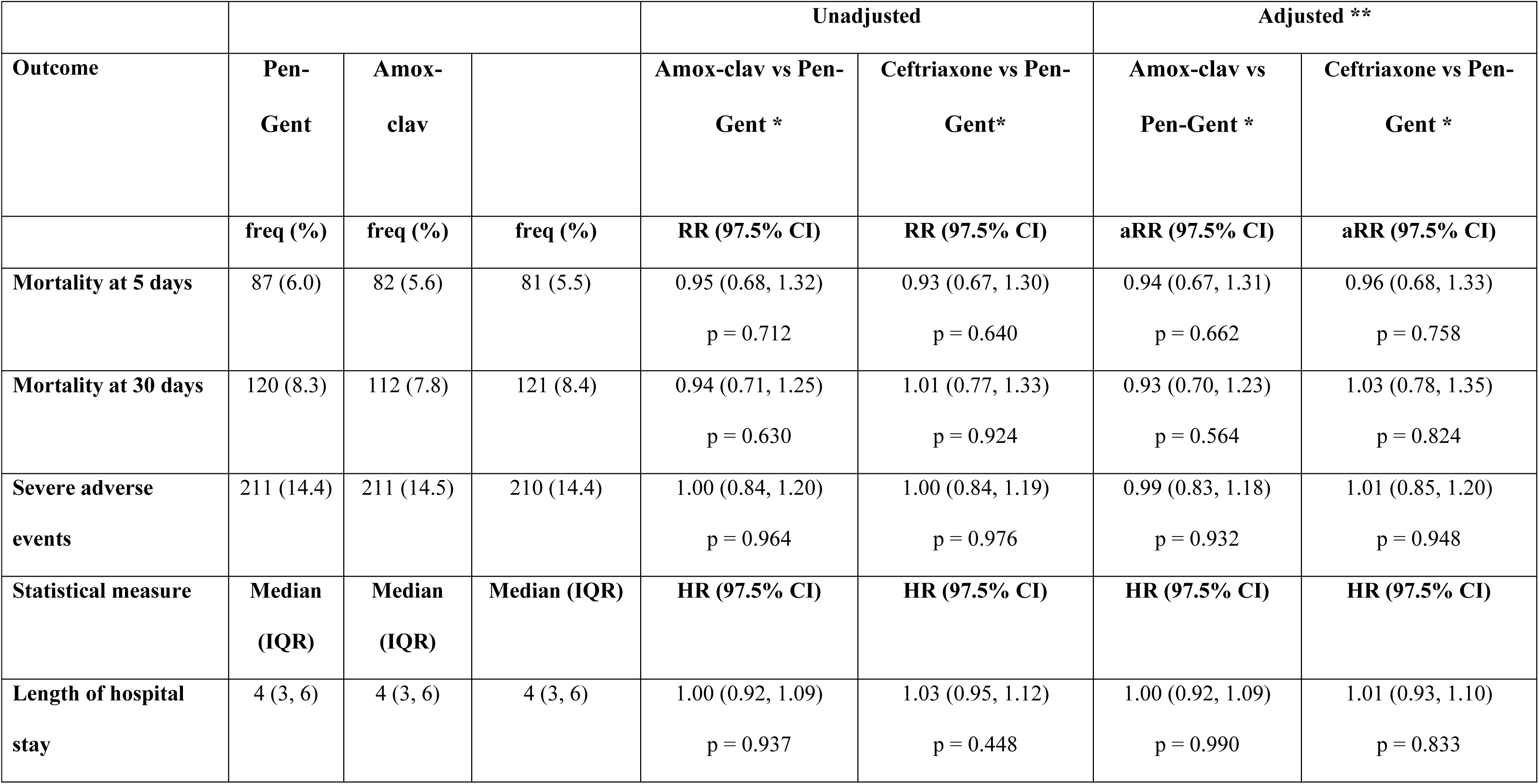
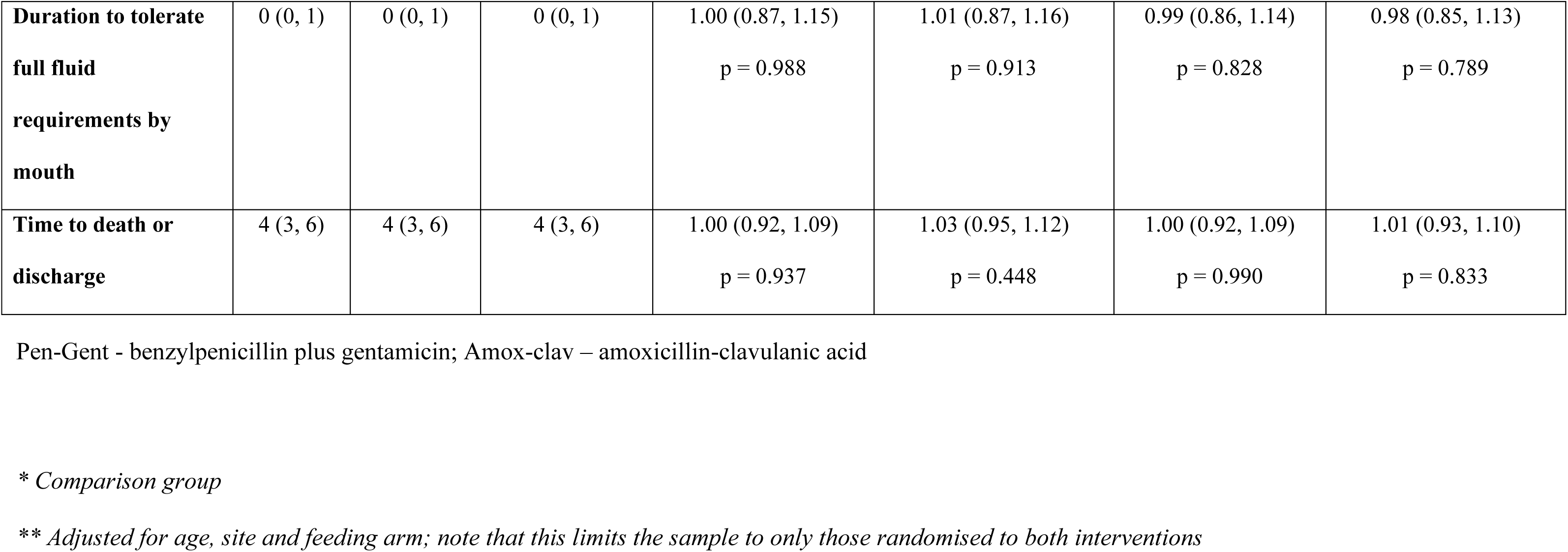
Primary and secondary outcomes by antibiotic treatment arm – Factorial Analysis.

|  |  |  |  | Unadjusted |  | Adjusted ** |  |
| --- | --- | --- | --- | --- | --- | --- | --- |
| Outcome | Pen-Gent | Amox-clav |  | Amox-clav vs Pen-Gent * | Ceftriaxone vs Pen-Gent* | Amox-clav vs Pen-Gent * | Ceftriaxone vs Pen-Gent * |
|  | freq (%) | freq (%) | freq (%) | RR (97.5% CI) | RR (97.5% CI) | aRR (97.5% CI) | aRR (97.5% CI) |
| Mortality at 5 days | 87 (6.0) | 82 (5.6) | 81 (5.5) | 0.95 (0.68, 1.32)<br>p = 0.712 | 0.93 (0.67, 1.30)<br>p = 0.640 | 0.94 (0.67, 1.31)<br>p = 0.662 | 0.96 (0.68, 1.33)<br>p = 0.758 |
| Mortality at 30 days | 120 (8.3) | 112 (7.8) | 121 (8.4) | 0.94 (0.71, 1.25)<br>p = 0.630 | 1.01 (0.77, 1.33)<br>p = 0.924 | 0.93 (0.70, 1.23)<br>p = 0.564 | 1.03 (0.78, 1.35)<br>p = 0.824 |
| Severe adverse events | 211 (14.4) | 211 (14.5) | 210 (14.4) | 1.00 (0.84, 1.20)<br>p = 0.964 | 1.00 (0.84, 1.19)<br>p = 0.976 | 0.99 (0.83, 1.18)<br>p = 0.932 | 1.01 (0.85, 1.20)<br>p = 0.948 |
| Statistical measure | Median (IQR) | Median (IQR) | Median (IQR) | HR (97.5% CI) | HR (97.5% CI) | HR (97.5% CI) | HR (97.5% CI) |
| Length of hospital stay | 4 (3, 6) | 4 (3, 6) | 4 (3, 6) | 1.00 (0.92, 1.09)<br>p = 0.937 | 1.03 (0.95, 1.12)<br>p = 0.448 | 1.00 (0.92, 1.09)<br>p = 0.990 | 1.01 (0.93, 1.10)<br>p = 0.833 |
| **Duration to tolerate full fluid requirements by mouth** | 0 (0, 1) | 0 (0, 1) | 0 (0, 1) | 1.00 (0.87, 1.15)<br>p = 0.988 | 1.01 (0.87, 1.16)<br>p = 0.913 | 0.99 (0.86, 1.14)<br>p = 0.828 | 0.98 (0.85, 1.13)<br>p = 0.789 |
| **Time to death or discharge** | 4 (3, 6) | 4 (3, 6) | 4 (3, 6) | 1.00 (0.92, 1.09)<br>p = 0.937 | 1.03 (0.95, 1.12)<br>p = 0.448 | 1.00 (0.92, 1.09)<br>p = 0.990 | 1.01 (0.93, 1.10)<br>p = 0.833 |

Pen-Gent - benzylpenicillin plus gentamicin; Amox-clav – amoxicillin-clavulanic acid
*\* Comparison group*
*\*\* Adjusted for age, site and feeding arm; note that this limits the sample to only those randomised to both interventions*

**Table 2b.**
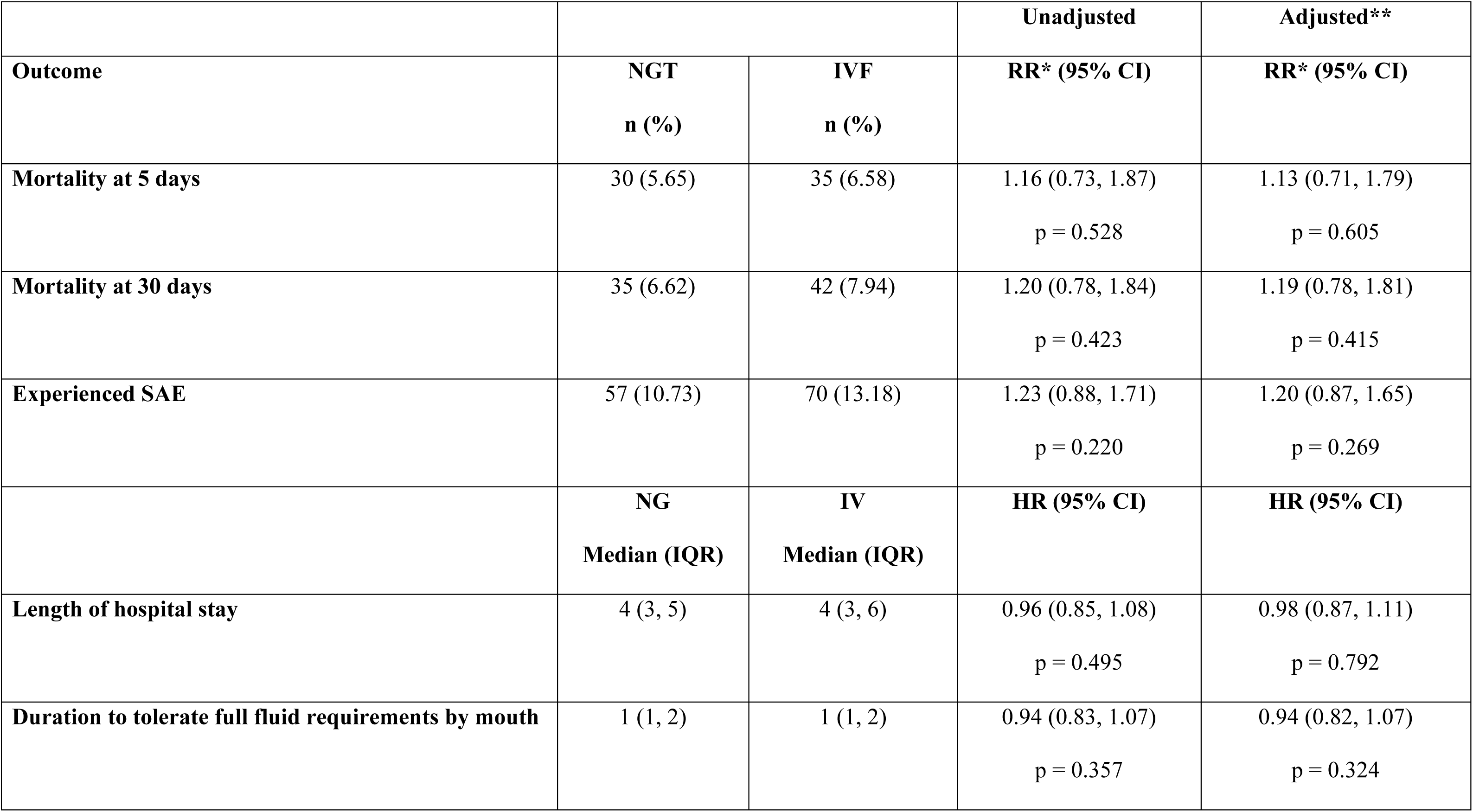
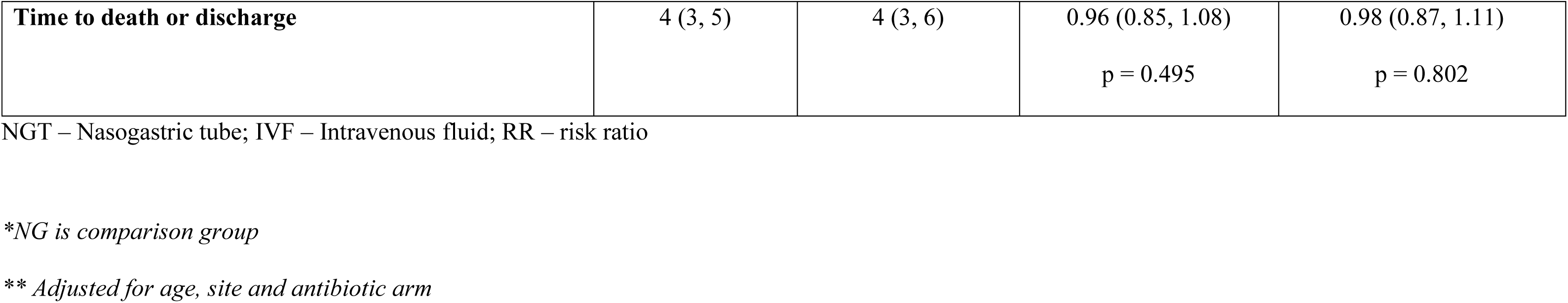
Primary and secondary outcomes by supportive care treatment arm – Factorial Analysis.

**Table 3.** Antibiotic and supportive care treatment non-compliance and reasons.

|  | Antibiotic |  |  | Supportive Care |  |
| --- | --- | --- | --- | --- | --- |
|  | **Pen-Gent**<br>**N = 237**<br>**n (%)** | **Amox-clav**<br>**N = 177**<br>**n (%)** | **Ceftriaxone**<br>**N = 41**<br>**n (%)** | **NGT**<br>**N = 312**<br>**n (%)** | **IVF**<br>**N = 271**<br>**n (%)** |
| **Changed** |  |  |  |  |  |
| **Reason\*** |  |  |  |  |  |
| Change of diagnosis | 29 (12.2) | 20 (11.3) | 3 (7.3) | 2 (0.6) | 1 (0.4) |
| Treatment failure | 24 (10.1) | 12 (6.8) | 1 (2.4) | 1 (0.3) | 2 (0.7) |
| Senior review | 168 (70.9) | 134 (75.7) | 30 (73.2) | 0 (0.0) | 2 (0.7) |
| Consent withdrawn | 3 (1.3) | 1 (0.6) | 3 (7.3) | 0 (0.0) | 1 (0.4) |
| Other | 17 (7.2) | 11 (6.2) | 4 (9.8) | 309 (99.0)** | 265 (97.8)** |
| *\* More than one may be specified*<br>*\*\* Other reason indicated as “able to tolerate feeds orally”* |  |  |  |  |  |

Pen-Gent - benzylpenicillin plus gentamicin; Amox-clav – amoxicillin clavulanic acid; NGT
– Nasogastric tube; IVF – Intravenous fluid

In the supportive-care comparison, deaths by day 5 occurred in 30/531 participants (5.7%) assigned to nasogastric feeding and 35/532 (6.6%) assigned to intravenous fluids (aRR, 1.13; 95% CI, 0.71 to 1.79) (Table 2B).

Prespecified subgroup analyses showed no evidence of heterogeneity of treatment effect according to HIV status, nutritional status, age, malaria status, or pallor (Figure 3a-3b). Post hoc analyses showed no variation in treatment effects across trial sites (Figures S4a-S4d).

**Figure 3a:**
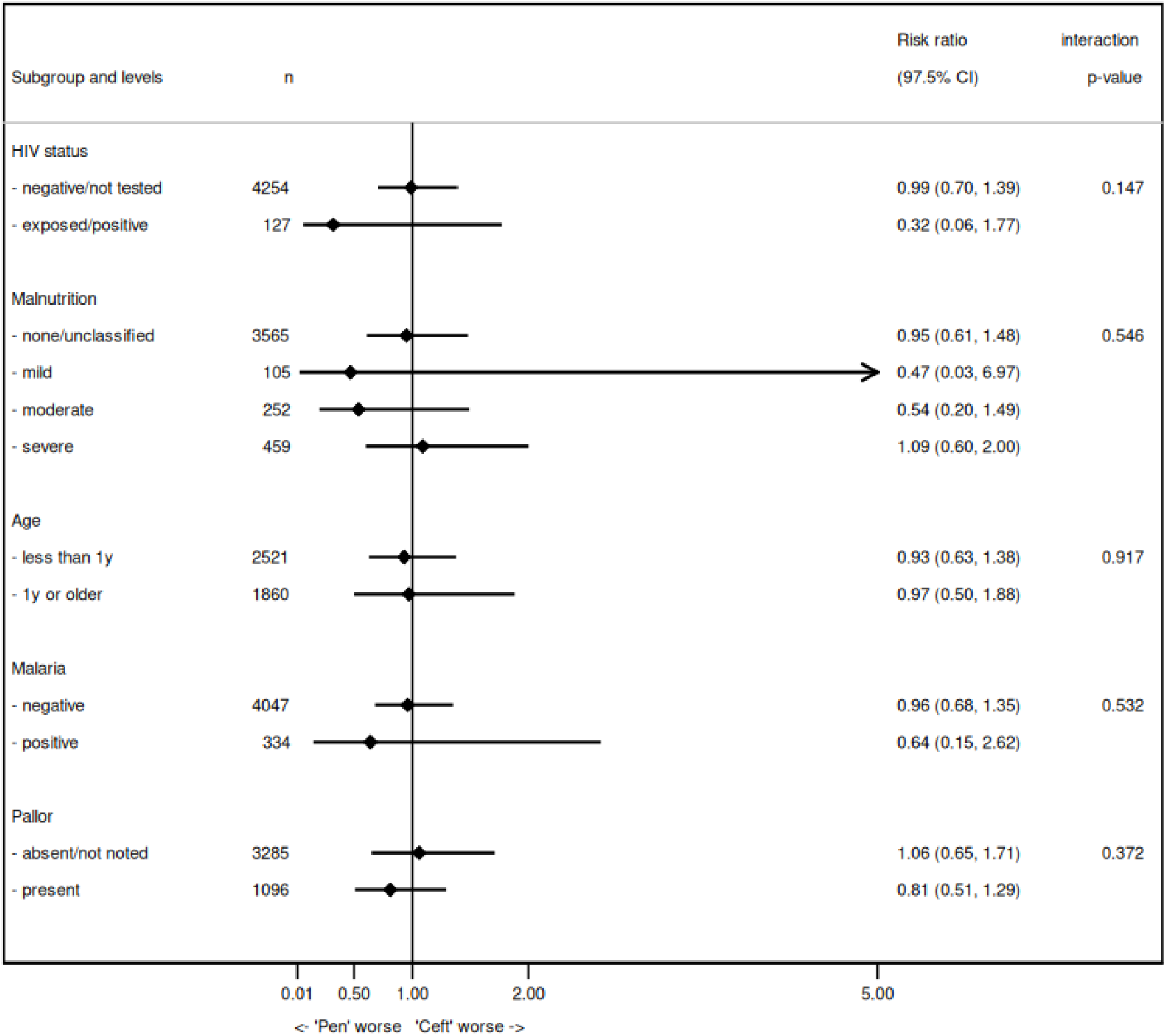
Subgroup analyses for day 5 mortality: Benzylpenicillin plus gentamicin vs Ceftriaxone

**Figure 3b:**
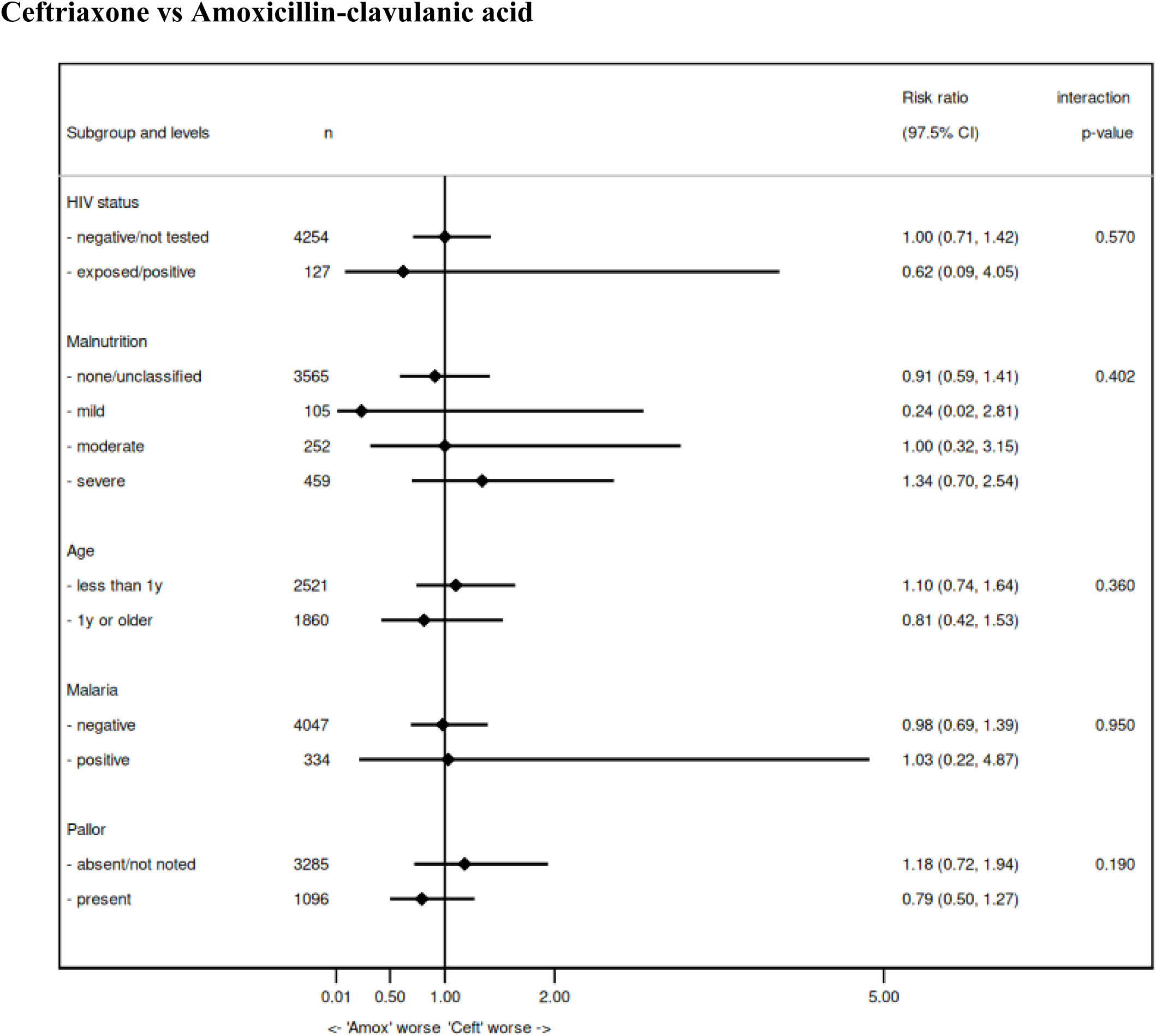
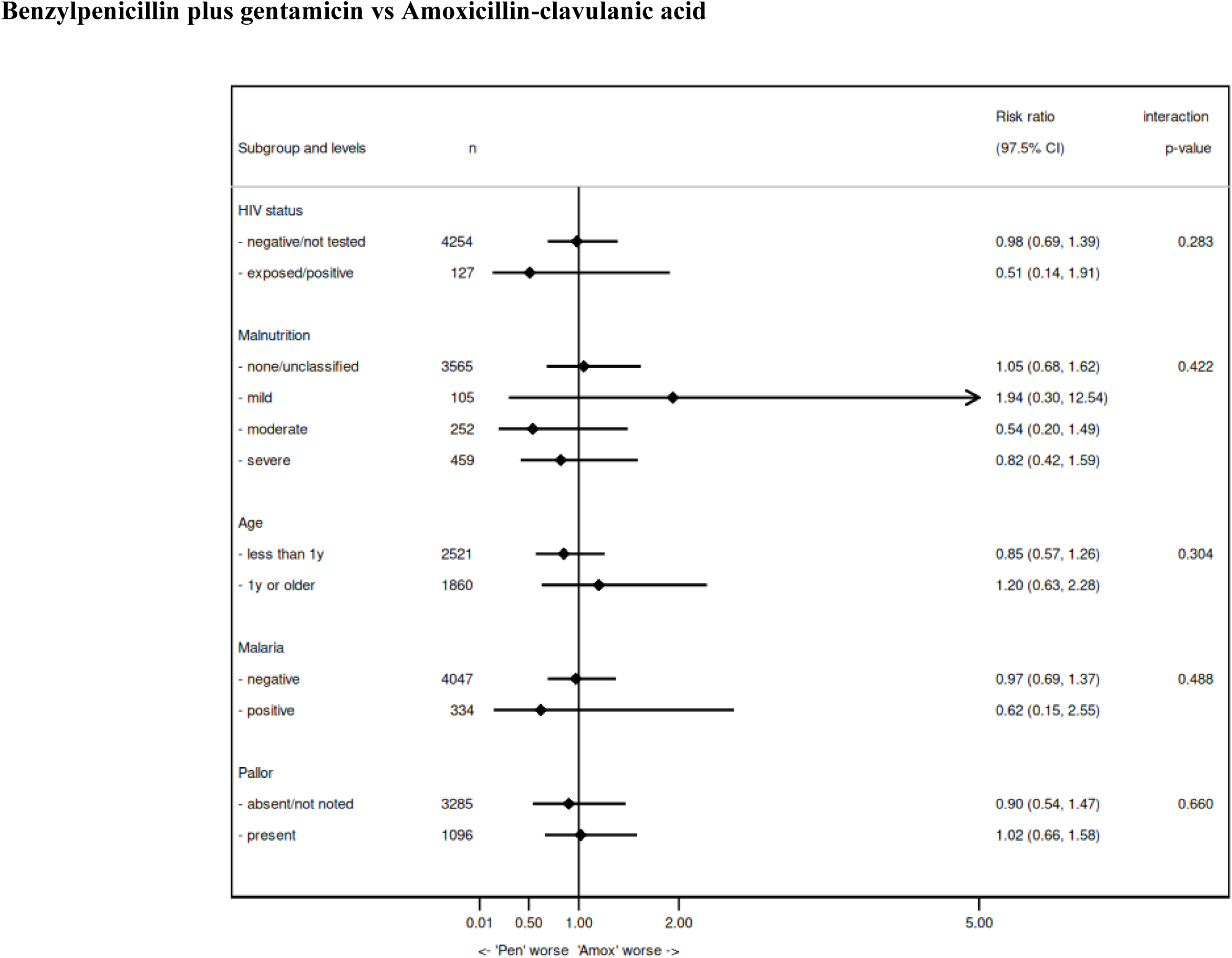
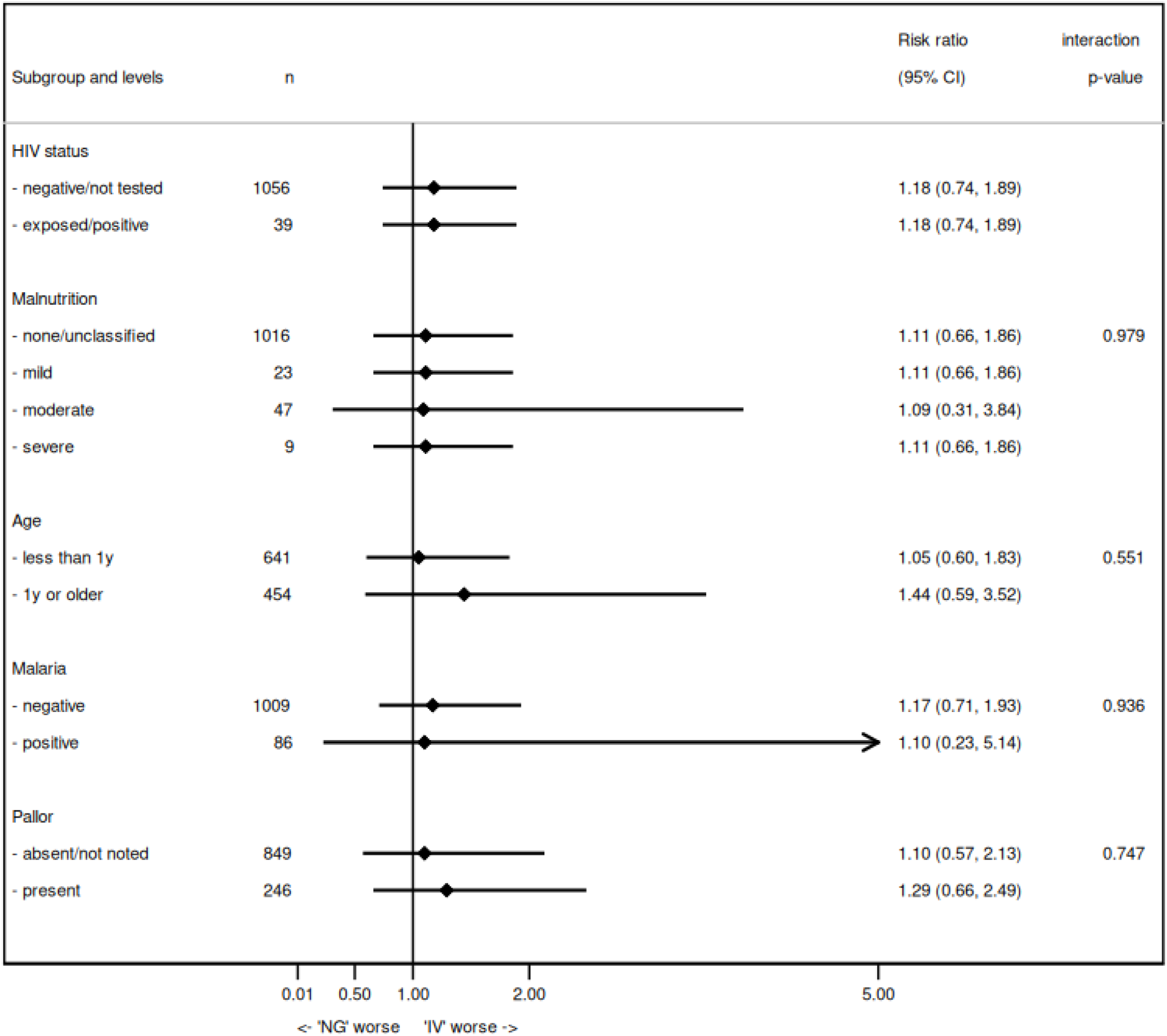
Subgroup analyses for day 5 mortality: Nasogastric tube feeding vs Intravenous fluid therapy

### Secondary Outcomes

By day 30, mortality remained similar across antibiotic groups: 120/1,445 participants (8.3%) in the benzylpenicillin plus gentamicin group, 112/1,445 (7.8%) in the amoxicillin-clavulanic acid group (aRR, 0.93; 95% CI, 0.70 to 1.23), and 121/1,455 (8.3%) in the ceftriaxone group (aRR, 1.02; 95% CI, 0.78 to 1.35) (Table 2A). In an exploratory comparison between ceftriaxone and amoxicillin-clavulanic acid, mortality was similar at day 5 (aRR 1.02; 95% CI, 0.73 to 1.43) and at day 30 (aRR 1.10; 95% CI, 0.83 to 1.46).

In the supportive-care comparison, 35/529 participants (6.6%) assigned to nasogastric feeding and 42/529 (7.9%) assigned to intravenous fluids had died by day 30 (aRR, 1.19; 95% CI, 0.79 to 1.81) (Table 2B).

Median length of hospital stay was 4 days (interquartile range, 3 to 6) across all groups. Time to full oral intake and time to death or discharge were also similar between groups (Tables 2A and 2B, and Figures 2A and 2B).

### Treatment Changes and Adherence

Changes in antibiotic therapy occurred more frequently among participants assigned to benzylpenicillin plus gentamicin (237 of 1,466; 16.2%) and amoxicillin–clavulanic acid (177 of 1,462; 12.1%) than among those assigned to ceftriaxone (41 of 1,465; 2.8%). In all groups, the most common reason for treatment change was clinical reassessment by a senior clinician (Table 4).

In the supportive care comparison, changes in feeding method were primarily attributable to clinical improvement and the ability to tolerate oral intake (Table 4).

Per-protocol analyses and complier-average causal-effect analyses yielded results consistent with the intention-to-treat analyses (Tables S3A through S3C).

### Safety

Serious adverse events (SAEs) occurred in 241 participants (16.4%) in the benzylpenicillin plus gentamicin group, 231 (15.8%) in the amoxicillin-clavulanic acid group, and 246 (16.8%) in the ceftriaxone group. In the supportive care comparison, SAEs occurred in 63 participants (11.8%) assigned to nasogastric feeding and 80 (15.0%) assigned to intravenous fluids. None of the SAEs were judged to be related to the study interventions.

## DISCUSSION

In this pragmatic, randomized trial involving children hospitalized with severe pneumonia, we found no meaningful differences in mortality or secondary clinical outcomes among those treated with benzylpenicillin plus gentamicin, ceftriaxone, or amoxicillin-clavulanic acid. Similarly, nasogastric tube feeding resulted in outcomes comparable to those with intravenous fluid therapy. These findings suggest that the WHO-recommended first-line antibiotic regimen remains appropriate for most children with severe pneumonia and that nasogastric feeding is a safe alternative to intravenous fluids for supportive care.

To our knowledge, this trial is the largest individually randomized evaluation of injectable antibiotic regimens in children hospitalized with severe pneumonia. Recommendations for severe pneumonia management (6) have remained largely unchanged for more than a decade and are based primarily on trials conducted more than two decades ago (32,33). Our findings provide contemporary evidence from routine-care settings that supports continued use of benzylpenicillin plus gentamicin as first-line therapy, without evidence of benefit from more costly, broader-spectrum alternatives. These results are particularly relevant in the context of increasing antimicrobial resistance, which is estimated to cause more than one million deaths annually worldwide and disproportionately affects low- and middle-income countries. (34)

The absence of detectable differences between antibiotic regimens likely reflects the changing epidemiology of childhood pneumonia, in which viral pathogens now predominate following widespread vaccination against Haemophilus influenzae type b and pneumococcus (2). In addition, prehospital antibiotic exposure was common in this population, reported in more than 80% of participants, consistent with findings from other low- and middle-income countries (35–39) Many of these prescriptions are driven by syndromic clinical algorithms that do not distinguish between bacterial and viral etiologies potentially diminishing detectable differences between antibiotic regimens. These findings highlight the limitations of symptom-based treatment approaches and underscore the need for improved diagnostic strategies to guide antibiotic use and strengthen antimicrobial stewardship (39–41).

This trial also represents the largest randomized evaluation of supportive care modalities in children with severe pneumonia. Although we observed no statistically significant differences between nasogastric feeding and intravenous fluid therapy, point estimates for mortality and recovery outcomes consistently favored nasogastric feeding, with no evidence of harm across subgroups. Nasogastric feeding provides both hydration and early nutritional support, preserves gastrointestinal function, and may avoid complications associated with intravenous access and fluid overload. Our findings are broadly consistent with a Cochrane review, which found no significant difference between enteral and intravenous rehydration in young children with bronchiolitis (42) based on findings from two RCTs conducted in Australia and New Zealand (43) and Israel (44). More recently, a trial involving children in four sub-Saharan African countries with severe acute malnutrition and gastroenteritis similarly showed no evidence of a mortality difference between oral and intravenous rehydration strategies, (45) reinforcing the safety of enteral approaches in critically ill children in low-resource settings. Together, these data suggest that enteral hydration and feeding strategies may be appropriate across a range of severe childhood illnesses when oral intake is not possible. If confirmed in larger pneumonia-specific trials, nasogastric feeding could represent an effective and pragmatic alternative to intravenous therapy, particularly in resource-limited settings where intravenous access may be challenging.

This study has several strengths. The trial enrolled a large number of participants across geographically diverse hospitals, enhancing generalizability to other low- and middle-income country settings. The pragmatic design ensured broad eligibility criteria and delivery of interventions within routine clinical workflows, increasing external validity. Adherence to assigned interventions was high, loss to follow-up was minimal, and the primary outcome of mortality was objective, reducing the risk of measurement bias. Consistent findings across intention-to-treat and per-protocol analyses further support the robustness of the results.

The trial also had limitations. Observed mortality was lower than anticipated, which reduced statistical power to detect modest differences between antibiotic regimens. A possible explanation is improved quality of care during trial participation; however, evidence for a consistent ‘trial effect’ is mixed and remains contentious (46) In addition, the supportive care comparison enrolled fewer participants than planned and was therefore underpowered to detect small differences. Finally, microbiological testing was not routinely performed, limiting our ability to relate treatment effects to pathogen-specific etiologies.

In conclusion, among children hospitalized with severe pneumonia, outcomes with benzylpenicillin plus gentamicin were similar to those with ceftriaxone or amoxicillin-clavulanic acid, supporting continued use of guideline-recommended first-line therapy. Nasogastric feeding resulted in outcomes comparable to those with intravenous fluid therapy and represents a feasible supportive care alternative in routine practice. These findings provide contemporary evidence to inform treatment guidelines and antimicrobial stewardship in low-resource settings and demonstrate the feasibility of conducting large pragmatic trials embedded within routine health systems to generate policy-relevant evidence.

## Declarations

### Author contributions

AA conceptualised the study with input from ME, EMO, and EA. LI, LM, JN, EMO, ME, EA, and AA jointly contributed to the study design. LI, LM, TN, JS, DK, RN, CW, MS, EI, EJ, AA, JB, MI, RI, AI, DL, FM, RM, CNM, JM, MM, CM, PM, EN, MN, CN, SO, LO, LT, MK, NM, EN, and AA supported the conduct of the trial. LI, CO, LM, EA, and AA contributed to data analysis and interpretation. LI drafted the initial manuscript with input from CO and AA. All authors critically reviewed, revised, and approved the final manuscript.

## Data Availability

Data requests will be considered by applying to the Data Governance Committee at CGMR-C Kilifi who will manage the process and ensure that appropriate ethical approval is in place and consent has been obtained for uses outside those of the original scope of work.

## Acknowledgments

We are grateful to all caregivers and participants. We would like to thank the Ministry of Health and county health management teams for their support. We would like to thank hospital teams including paediatric ward staff, pharmacy staff, health records and information department staff, hospital administration. We would like to thank all study clinicians and data clerks : David Mugambi, Sylvia Namu, Bancy Mumbi, Joseph Njoroge, Eva Kamau, Mary Waithera, Joy-Patience Murugi, Kennedy Kinyua, Protus Changaya, Kivuli Ayudi, Felistus Mumbi, Geoffrey Oeri, Perpetua Mule, Shukri Juweiriya, Grace Atieno, Irene Maina, Edwin Ryan Otieno, Janet Mogendi, Elkanah Sitienei, Alex Kaara, Zachariah Kiama, Lynn Mwendwa, Lilian Wairimu Muhoro, Harun Kuria, Kevin Kiruki, Kevin Okumu, Rose Murere, Jane Kenani, Jemimah Jesse, Wilfred Masososo, Mercy Manyonge, Jedidah Songwe, Andrew Wanyonyi, Peter Omondi, Selestine Adhiambo, Collins Ekisa, Peter Wandera, Abraham Wahanga, Beason Soita, Denish Ouma Otieno, David Simiyu, Jackson Obare, Brenda Wafula, Benjamin Ayiecha, Sharon Auma, Meshack Tumwa, Adnan Odondi, Mark Robert Obuya, Millicent Ogollo, James Aloo, Kennedy Otiende, Damaris Chepkemoi, Cecilia Agutu and Dennis Wafula. We would like to thank the hospital pharmacists Dr Carolyne Naliaka, Dr Sharon Apinde, Dr Esther Ngethe, Dr Linda Achieng, Dr Lindsay Olima, Dr Cynthia Nduta, Dr Rodgers Omollo, Dr Steve Gichana Makori, Dr Evans Gitu, Dr Evangeline Maina, Dr Evans Makumba, Dr Neha Bilakhia, Judy Wanjiru Kariuki, Dr Francis Wachuri, Dr Lilian Kabesa and Dr Judy Odenyo.

We gratefully acknowledge the invaluable guidance and oversight provided by the independent Trial Steering Committee members: Prof. Richard Lilford (Chair), Prof. Peter Waiswa, and Dr. Jane Crawley. We also thank the Data and Safety Monitoring Board members: Prof. Festus Kalokola (Chair), Dr. Isaac Tsikhustsu, and Prof. Andrew Copas.

We also acknowledge the UK Department of Health and Social Care, the UK Foreign Commonwealth & Development Office, the UK Medical Research Council, and the Wellcome Trust through the Joint Global Health Trials Scheme.

## Participant recruitment and retention

## Notes

### Competing Interest Statement

The authors have declared no competing interest.

### Clinical Trial

NCT04041791

### Funding Statement

This study was supported by the UK Department of Health and Social Care, the UK Foreign, Commonwealth & Development Office, the UK Medical Research Council, and Wellcome through the Joint Global Health Trials Scheme. The funders provided financial support for the conduct of the trial. The authors and their institutions did not receive any additional payment or services from a third party for study design, data analysis, manuscript preparation, or the decision to submit for publication. The funders had no role in the design of the study, collection, analysis, or interpretation of the data, preparation of the manuscript, or the decision to submit for publication.

### Author Declarations

The Scientific and Ethics Review Unit of the Kenya Medical Research Institute, the Pharmacy and Poisons Board of Kenya, the National Commission for Science, Technology and Innovation of Kenya, and the Oxford Tropical Research Ethics Committee of the University of Oxford gave ethical approval for this work.

### Summary of Updates

In this revised version, the funding statement has been updated to provide the full names of all supporting organizations. No changes were made to the study data, analyses, or conclusions.

