## Supplementary Appendix for "A Pragmatic Trial of Antibiotics and Supportive Care for Severe Pneumonia in Hospitalized Children"

### Supportive Care and Antibiotics for Severe Pneumonia among Hospitalised Children: A Pragmatic Randomized Controlled Trial.

**List of Investigators**

Lynda Isaaka,[1], Charles Opondo, PhD[2], Livingstone Mumelo BSc[1], Teresiah Njoroge, MMed[1], Jimmy Shangala, MSc[1], Dennis Kimego BSc[1], Rebecca Njuguna, MSc[1], Conrad Wanyama MSc[1], Metrine Saisi MSc[1], Elizabeth Isinde BSc[1], Elizabeth Jowi, MMed [3], Achieng Adem, MMed [4], Julie Barasa, MMed [5], Mourine Ikol, MMed [6], Rachael Inginia, MMed [7], Angeline Ithondeka, MMed [5,17], Dickens Lubanga, MMed [8], Felicitas Makokha, MMed [8], Roselyn Malangachi, MMed [9], Christine Marete, MMed [10], Jecinter Modi, MMed [11], Maureen Muchela, MMed [12], Celia Muturi, MMed [3], Peninnah Mwangi, MMed [13], Emma Namulala, MMed [14], Maureen Njoroge, MMed [15], Charles Nzioki, MMed [16], Sharon Ocharo, MMed [4], Linda Ombito, MMed [17], Lydia Thuranira ,MMed [18], Magdaline Kuria, MMed [4], Ngina Mwangi, MMed [10], Esther Njiru, MMed [10], James Nokes, PhD [1], Grace Irimu, PhD [1] , Frederick N Were, PhD [19], Sam Akech, PhD [1], Edwine Barasa, PhD [1], Elizabeth Maleche Obimbo, PhD [21], Mike English, PhD [20], Elizabeth Allen, PhD [2], Ambrose Agweyu, PhD [1,2]

[1]KEMRI-Wellcome Trust Research Programme, Kenya

[2]London School of Hygiene and Tropical Medicine, London, United Kingdom.

[3]Mama Lucy Kibaki Hospital, Nairobi, Kenya.

[4]Kisumu County Referral Hospital, Kisumu, Kenya.

[5]Naivasha County Referral Hospital, Naivasha, Kenya.

[6]Kisii Teaching and Referral Hospital, Kisii, Kenya

[7]Kitale County Referral Hospital, Kitale, Kenya.

[8]Bungoma County Referral Hospital, Bungoma, Kenya.

[9]Kakamega County General Hospital, Kakamega, Kenya.

[10]Embu Level 5 Hospital, Embu, Kenya

[11]Kenyatta National Hospital, Nairobi, Kenya

[12]Jaramogi Oginga Odinga Teaching and Referral Hospital, Kisumu, Kenya

[13]Kerugoya County Referral Hospital, Kerugoya, Kenya.

[14]Busia County Referral Hospital, Busia, Kenya.

[15]Thika Level Five Hospital, Thika, Kenya.

[16]Machakos Level Five Hospital, Kenya.

[17]Nakuru Level Six Hospital, Nakuru, Kenya.

[18]Kiambu Country Referral Hospital, Kiambu,  Kenya.

[19]Kenya Paediatric Research Consortium, Kenya

[20]University of Oxford, Oxford, United Kingdom.

[21]University of Nairobi, Nairobi, Kenya.

**Table S1: WHO 2013 Guidelines for Classification and Management of Severe Pneumonia**(1)

The table below summarises the WHO criteria for the classification of pneumonia in children aged 2-59 years.

| Syndrome | Clinical signs in children aged 2-59 months with cough and/or difficulty in breathing | Recommended setting for management and antibiotic treatment |
|---|---|---|
| **Severe pneumonia** | Any one of: oxygen saturation <90%, central cyanosis, inability to drink/breastfeed, vomiting everything, altered consciousness, convulsion | • Inpatient management<br>• Intravenous benzylpenicillin/ampicillin and gentamicin |
| **Non-severe pneumonia** | Lower chest wall indrawing<br>OR<br>Fast breathing (respiratory rate > 50/min if aged 2-11 months ; 40/min if aged 12-59 months)<br>AND<br>Without signs of severe pneumonia | • Outpatient management<br>• Oral amoxicillin |

**Table S2: Study Sites**

The SEARCH trial was conducted in 16 hospitals across Kenya that were part of a clinical information network.(2) These hospitals served diverse populations from urban, peri-urban and rural areas in Western and Central Kenya.

| | Study Site | Location |
|---|---|---|
| 1. | Embu Level 5 Hospital | Central Kenya; low malaria transmission |
| 2. | Kiambu County Referral Hospital | Central Kenya; low malaria transmission |
| 3. | Kerugoya County Referral Hospital | Central Kenya; low malaria transmission |
| 4. | Kenyatta National Hospital | Central Kenya; low malaria transmission |
| 5. | Machakos Level 5 Hospital | Central Kenya; low malaria transmission |
| 6. | Mama Lucy Kibaki Hospital | Central Kenya; low malaria transmission |
| 7. | Naivasha County Referral Hospital | Central Kenya; low malaria transmission |
| 8. | Nakuru Level 6 Hospital | Central Kenya; low malaria transmission |
| 9. | Thika Level 5 Hospital | Western Kenya ; high malaria transmission |
| 10. | Bungoma County Referral Hospital | Western Kenya ; high malaria transmission |
| 11. | Busia County Referral Hospital | Western Kenya ; high malaria transmission |
| 12. | Jaramogi Oginga Odinga Teaching and Referral Hospital | Western Kenya ; high malaria transmission |
| 13. | Kakamega Teaching and Referral Hospital | Western Kenya ; high malaria transmission |
| 14. | Kisii Teaching and Referral Hospital | Western Kenya ; high malaria transmission |
| 15. | Kisumu County Referral Hospital | Western Kenya ; high malaria transmission |
| 16. | Kitale County Referral Hospital | Western Kenya ; high malaria transmission |

**Figure S1: Factorial Randomisation Scheme**

The figure below summarises the participant randomisation scheme. All participants were randomly assigned to one of three antibiotics. Eligible participants were further randomised to receive either nasogastric feeds or intravenous fluids.

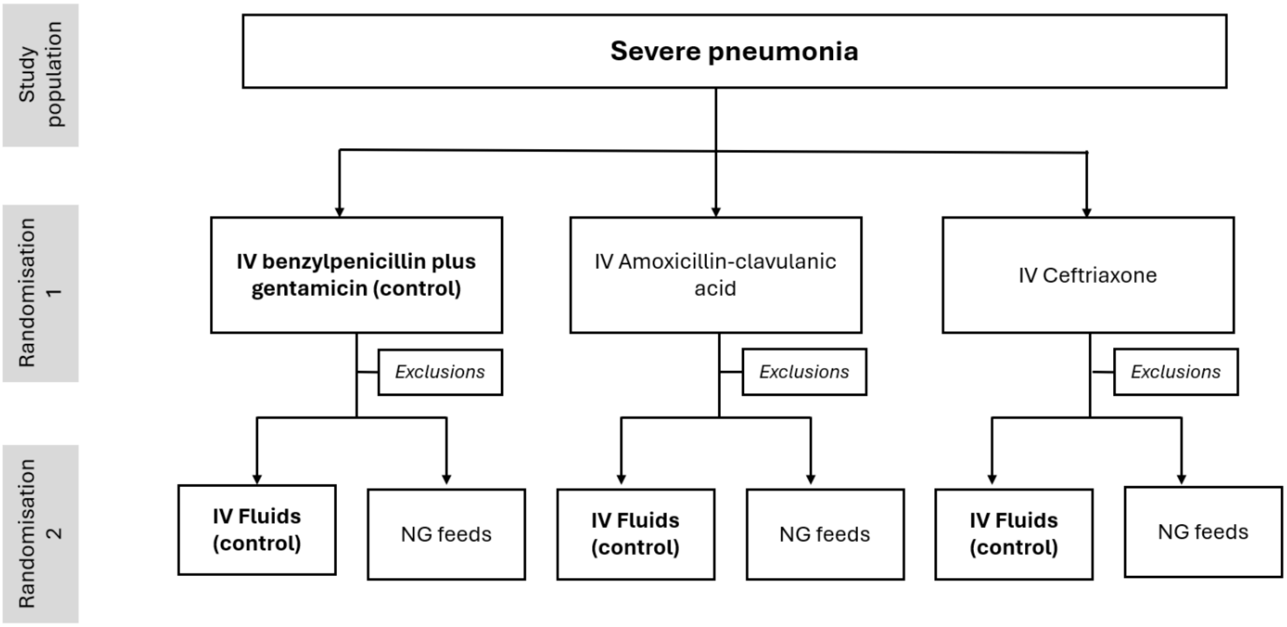

*IV -Intravenous* *NG- nasogastric*

**Table S3 Additional participant baseline characteristics by treatment arm**

| | | Antibiotic | | | Supportive Care | |
|---|---|---|---|---|---|---|
| **Characteristics** | **Total**<br>**N = 4,393** | **Pen-gent**<br>**N = 1,466** | **Amox-clav**<br>**N = 1,462** | **Ceft**<br>**N = 1,465** | **NGT**<br>**N = 532** | **IVF**<br>**N = 532** |
| **History and socio-demographic profile** | | | | | | |
| Age (months) – mean (SD) | 14 (12.3) | 14 (12.2) | 14 (12.2) | 14 (12.4) | 14 (2.6) | 13 (2.4) |
| WHZ – mean (SD) | 5 (1.4) | 5 (1.4) | 5 (1.5) | 5 (1.5) | 5 (0.9) | 5 (0.9) |
| MUAC (cm) (> 6months) – mean (SD) | 14 (2.6) | 14 (1.9) | 14 (3.6) | 14 (1.8) | 14 (2.7) | 14 (2.6) |
| Vomiting – Yes – n (%) | 1456 (33.1) | 496 (33.8) | 489 (33.4) | 471 (32.2) | 141 (26.5) | 148 (27.8) |
| Freq. of vomiting in 24 hrs– mean (SD) | 3 (1.8) | 3 (1.8) | 3 (1.7) | 3 (1.8) | 2 (0.4) | 2 (0.4) |
| Respiratory rate (bmp) – mean (SD) | 60 (13.2) | 60 (13.4) | 61 (12.9) | 60 (13.3) | 64 (12) | 64 (12) |
| O2 saturation (%)– mean (SD) | 87 (7.5) | 87 (7.4) | 87 (7.7) | 87 (7.5) | 87 (16.3) | 87 (16.3) |
| Central Cyanosis – n (%) | 55 (1.3) | 15 (1) | 21 (1.4) | 19 (1.3) | 9 (1.7) | 11 (2.1) |
| Grunting – n (%) | 2517 (57.3) | 806 (55) | 841 (57.5) | 870 (59.4) | 331 (62.2) | 367 (69) |
| Capillary refill (/secs) – n (%) | | | | | | |

| | | | | | | |
|---|---|---|---|---|---|---|
| 1 second | 3391 (77.2) | 1142 (77.9) | 1125 (76.9) | 1124 (76.7) | 431 (81) | 428 (80.5) |
| 2 seconds | 799 (18.2) | 259 (17.7) | 266 (18.2) | 274 (18.7) | 88 (16.5) | 92 (17.3) |
| 3+seconds | 199 (4.5) | 63 (4.3) | 71 (4.9) | 65 (4.4) | 13 (2.4) | 12 (2.3) |
| Unknown | 4 (0.1) | 2 (0.1) | 0 (0) | 2 (0.1) | 0 (0) | 0 (0) |
| Pallor – n (%) | | | | | | |
| 0 | 3288 (74.8) | 1094 (74.6) | 1105 (75.6) | 1089 (74.3) | 407 (76.5) | 413 (77.6) |
| + | 938 (21.4) | 319 (21.8) | 298 (20.4) | 321 (21.9) | 98 (18.4) | 99 (18.6) |
| +++ | 160 (3.6) | 50 (3.4) | 55 (3.8) | 55 (3.8) | 27 (5.1) | 18 (3.4) |
| Disability (AVPU) – n (%) | | | | | | |
| Alert | 4127 (93.9) | 1372 (93.6) | 1370 (93.7) | 1385 (94.5) | 471 (88.5) | 478 (89.8) |
| Responsive to voice | 173 (3.9) | 56 (3.8) | 61 (4.2) | 56 (3.8) | 55 (10.3) | 43 (8.1) |
| Responsive to pain | 80 (1.8) | 34 (2.3) | 26 (1.8) | 20 (1.4) | 5 (0.9) | 8 (1.5) |
| Unresponsive | 12 (0.3) | 4 (0.3) | 4 (0.3) | 4 (0.3) | 1 (0.2) | 3 (0.6) |
| Unknown | 1 (0) | 0 (0) | 1 (0.1) | 0 (0) | 0 (0) | 0 (0) |
| **Additional diagnoses at admission** | | | | | | |
| Malaria – n (%) | | | | | | |

| | | | | | | |
|---|---|---|---|---|---|---|
| Severe | 216 (4.9) | 75 (5.1) | 70 (4.8) | 71 (4.8) | 36 (6.8) | 29 (5.5) |
| Non-severe | 104 (2.4) | 37 (2.5) | 35 (2.4) | 32 (2.2) | 13 (2.4) | 12 (2.3) |
| Diarrhoea – n (%) | | | | | | |
| Non-bloody | 917 (20.9) | 295 (20.1) | 299 (20.5) | 323 (22) | 81 (15.2) | 78 (14.7) |
| Bloody | 16 (0.4) | 3 (0.2) | 6 (0.4) | 7 (0.5) | 0 (0) | 3 (0.6) |
| Not categorised | 5 (0.1) | 4 (0.3) | 1 (0.1) | 0 (0) | 1 (0.2) | 0 (0) |
| Dehydration – n (%) | | | | | | |
| Shock | 68 (1.5) | 23 (1.6) | 23 (1.6) | 22 (1.5) | 0 (0) | 0 (0) |
| Severe | 184 (4.2) | 57 (3.9) | 65 (4.4) | 62 (4.2) | 1 (0.2) | 1 (0.2) |
| Some | 299 (6.8) | 97 (6.6) | 100 (6.8) | 102 (7) | 25 (4.7) | 18 (3.4) |
| Not categorised | 3 (0.1) | 0 (0) | 1 (0.1) | 2 (0.1) | 0 (0) | 2 (0.4) |
| HIV – n (%) | | | | | | |
| Positive | 8 (0.2) | 3 (0.2) | 2 (0.1) | 3 (0.2) | 0 (0) | 0 (0) |
| Exposed / PMTCT+ | 119 (2.7) | 43 (2.9) | 44 (3) | 32 (2.2) | 16 (3) | 21 (3.9) |
| Malnutrition – n (%) | | | | | | |
| Mild | 119 (2.7) | 37 (2.5) | 41 (2.8) | 41 (2.8) | 12 (2.3) | 13 (2.4) |
| Moderate | 278 (6.3) | 93 (6.3) | 91 (6.2) | 94 (6.4) | 25 (4.7) | 29 (5.5) |
| Severe | 464 (10.6) | 151 (10.3) | 151 (10.3) | 162 (11.1) | 1 (0.2) | 4 (0.8) |
| Marasmus | 0 (0) | 0 (0) | 0 (0) | 0 (0) | 0 (0) | 0 (0) |
| Kwashiorkor | 4 (0.1) | 1 (0.1) | 3 (0.2) | 0 (0) | 0 (0) | 0 (0) |

| | | | | | | |
|---|---|---|---|---|---|---|
| Marasmus-Kwashiorkor | 11 (0.3) | 3 (0.2) | 4 (0.3) | 4 (0.3) | 0 (0) | 0 (0) |
| Anaemia – n (%) | | | | | | |
| Severe | 199 (4.5) | 65 (4.4) | 71 (4.9) | 63 (4.3) | 26 (4.9) | 21 (3.9) |
| Non-severe | 1603 (36.5) | 521 (35.5) | 539 (36.9) | 543 (37.1) | 196 (36.8) | 189 (35.5) |
| Not categorised | 3 (0.1) | 1 (0.1) | 2 (0.1) | 0 (0) | 0 (0) | 0 (0) |
| Asthma – n (%) | | | | | | |
| Very severe | 0 (0) | 0 (0) | 0 (0) | 0 (0) | 0 (0) | 0 (0) |
| Severe | 86 (2) | 26 (1.8) | 29 (2) | 31 (2.1) | 6 (1.1) | 15 (2.8) |
| Not severe | 66 (1.5) | 28 (1.9) | 15 (1) | 23 (1.6) | 7 (1.3) | 8 (1.5) |
| Not categorised | 7 (0.2) | 3 (0.2) | 3 (0.2) | 1 (0.1) | 1 (0.2) | 0 (0) |
| Suspected TB – n (%) | 93 (2.1) | 27 (1.8) | 39 (2.7) | 27 (1.8) | 8 (1.5) | 3 (0.6) |
| Sickle cell disease – n (%) | 66 (1.5) | 24 (1.6) | 18 (1.2) | 24 (1.6) | 10 (1.9) | 3 (0.6) |
| Meningitis – n (%) | 5 (0.1) | 2 (0.1) | 3 (0.2) | 0 (0) | 1 (0.2) | 0 (0) |
| Rickets – n (%) | 178 (4.1) | 58 (4) | 64 (4.4) | 56 (3.8) | 3 (0.6) | 4 (0.8) |
| Weight-for-height z-score | -1 (2.3) | -1 (1.8) | -1 (2.7) | -1 (2.4) | -1 (-0.1) | 0 (-0.1) |

**Note: Percentages are column percentages (i.e. overall and within-group prevalence).**

**Figure S2: Kaplan-Meier plot for length of hospital stay**

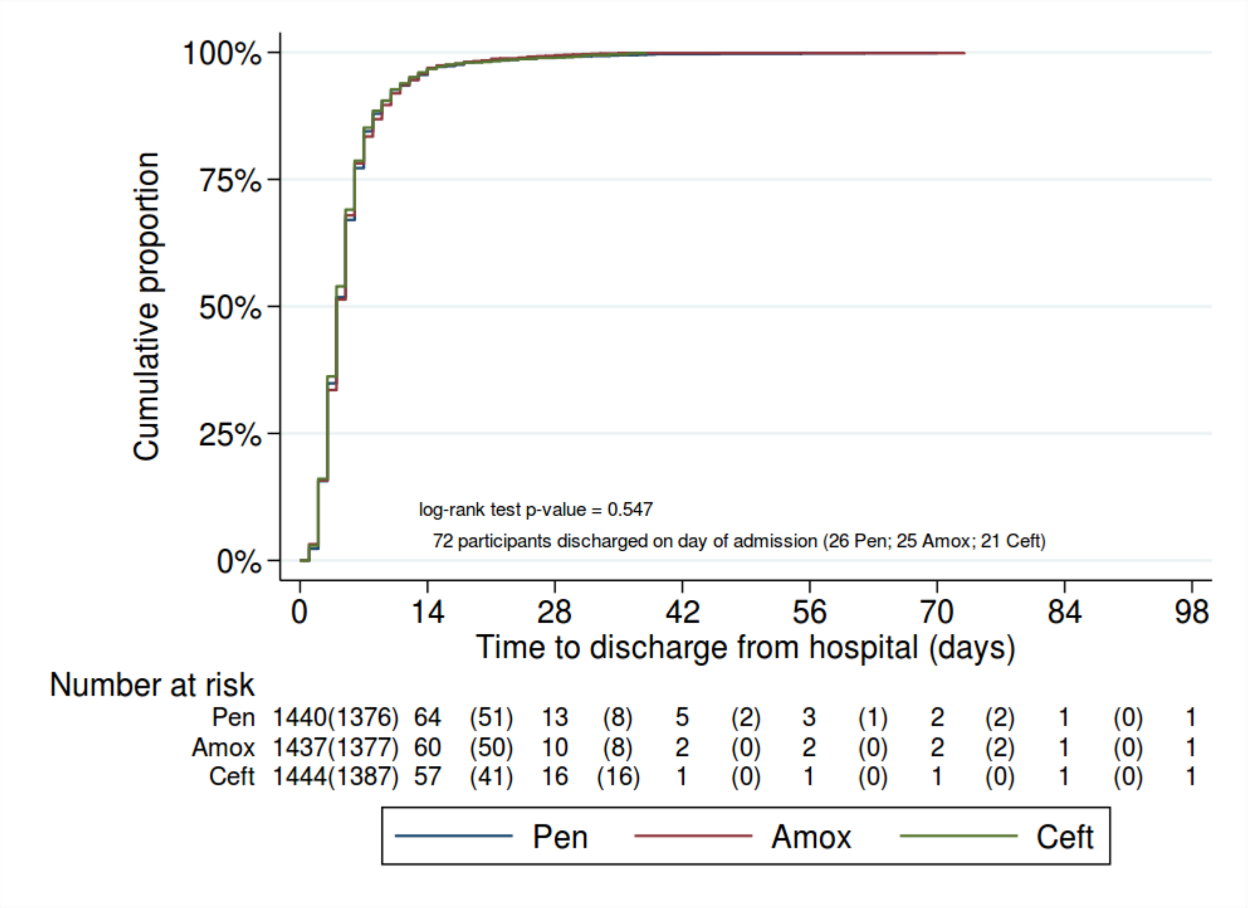

**Figure S3 Kaplan-Meier plot for duration to tolerate oral feeds**

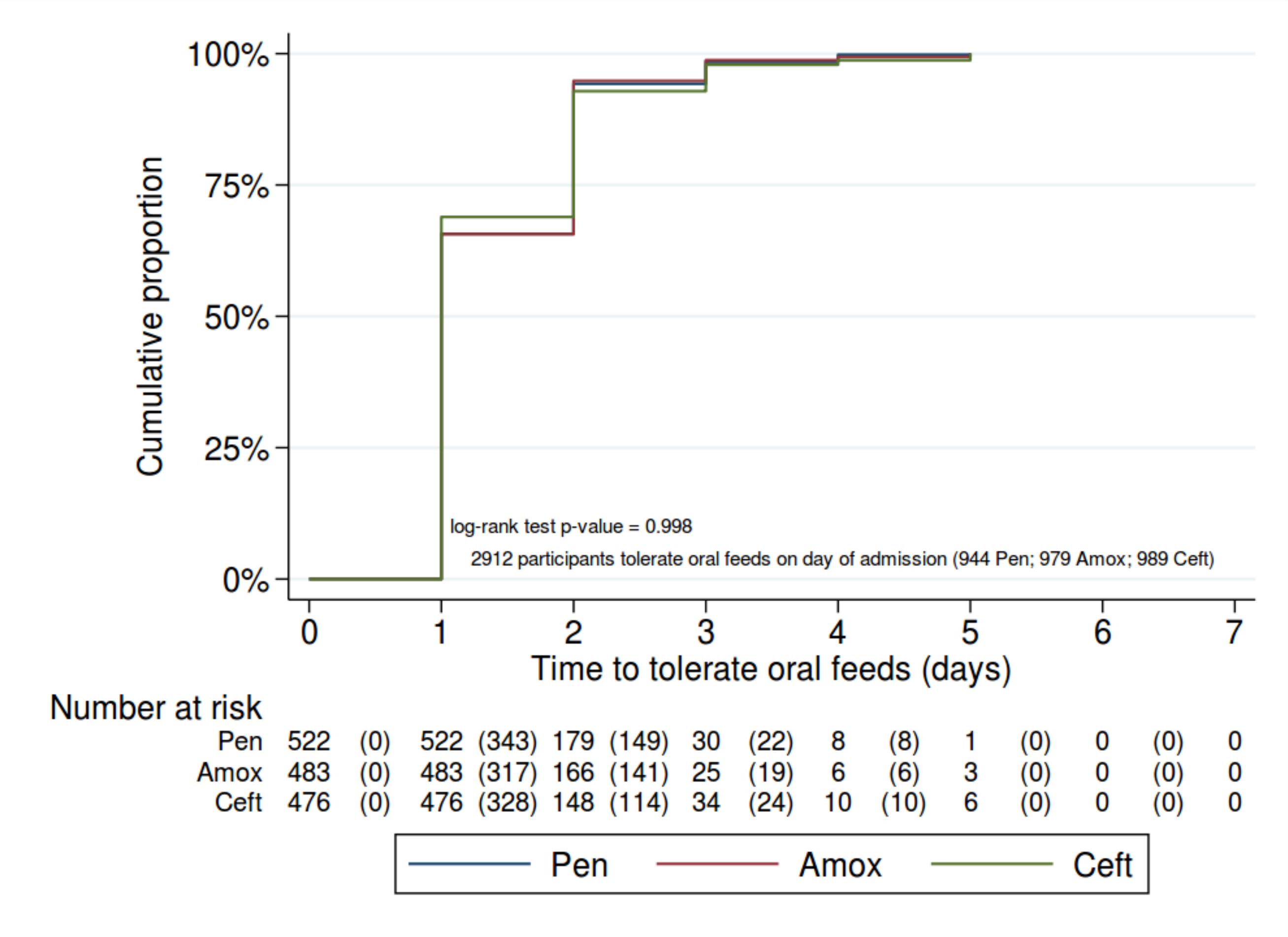

**Table S4a: Participants data flow by study site – Antibiotic Arm**

| Site [a] | Admitted N = 49341 | Eligible n (%) | Enrolled n (%) | Pen n | Amox n | Ceft n | Day 5 outcomes n (%) | Day 30 outcomes n (%) | Lost-to-follow-up day 5 n | Withdrew day 5 n | Lost-to-follow-up day 30 n | Withdrew day 30 n |
|---|---|---|---|---|---|---|---|---|---|---|---|---|
| **Antibiotic Treatment** | | | | | | | | | | | | |
| A | 203 | 203 (100) | 203 (100) | 68 | 68 | 67 | 199 (98) | 196 (96.6) | 0 | 3 | 2 | 0 |
| B | 2713 | 267 (9.8) | 220 (8.1) | 74 | 73 | 73 | 218 (99.1) | 211 (95.9) | 0 | 2 | 6 | 0 |
| C | 2833 | 172 (6.1) | 167 (5.9) | 57 | 55 | 55 | 167 (100) | 166 (99.4) | 0 | 0 | 1 | 0 |
| D | 3597 | 499 (13.9) | 396 (11) | 133 | 130 | 133 | 396 (100) | 396 (100) | 0 | 0 | 0 | 0 |
| E | 4169 | 436 (10.5) | 298 (7.1) | 99 | 98 | 101 | 297 (99.7) | 294 (98.7) | 0 | 1 | 3 | 0 |
| F | 1923 | 389 (20.2) | 300 (15.6) | 102 | 100 | 98 | 300 (100) | 299 (99.7) | 0 | 0 | 1 | 0 |
| G | 8545 | 1117 (13.1) | 690 (8.1) | 229 | 230 | 231 | 688 (99.7) | 678 (98.3) | 0 | 2 | 9 | 0 |
| H | 363 | 26 (7.2) | 22 (6.1) | 8 | 7 | 7 | 22 (100) | 21 (95.5) | 0 | 0 | 1 | 0 |
| I | 2405 | 305 (12.7) | 294 (12.2) | 99 | 97 | 98 | 294 (100) | 293 (99.7) | 0 | 0 | 1 | 0 |
| J | 2352 | 307 (13.1) | 302 (12.8) | 101 | 100 | 101 | 302 (100) | 301 (99.7) | 0 | 0 | 1 | 0 |
| K | 4912 | 364 (7.4) | 347 (7.1) | 115 | 118 | 114 | 345 (99.4) | 341 (98.3) | 0 | 2 | 4 | 0 |
| L | 4495 | 349 (7.8) | 340 (7.6) | 113 | 113 | 114 | 340 (100) | 338 (99.4) | 0 | 0 | 2 | 0 |
| M | 2957 | 246 (8.3) | 203 (6.9) | 66 | 69 | 68 | 202 (99.5) | 197 (97) | 0 | 1 | 5 | 0 |
| N | 6609 | 529 (8) | 519 (7.9) | 173 | 173 | 173 | 519 (100) | 515 (99.2) | 0 | 0 | 4 | 0 |
| O | 657 | 54 (8.2) | 50 (7.6) | 16 | 17 | 17 | 50 (100) | 50 (100) | 0 | 0 | 0 | 0 |
| P | 608 | 51 (8.4) | 42 (6.9) | 13 | 14 | 15 | 42 (100) | 42 (100) | 0 | 0 | 0 | 0 |
| **Total** | **49341** | **5314 (10.8)** | **4393 (8.9)** | **1466** | **1462** | **1465** | **4381 (99.7)** | **4338 (98.7)** | **0** | **11** | **40** | **0** |

**Table S4b: Participants data flow by study site – Supportive Care Arm**

| Site | Admitted N = 49341 | Eligible n (%) | Enrolled n (%) | NG n | IV n | | Day 5 outcomes n (%) | Day 30 outcomes n (%) | Lost-to-follow-up day 5 n | Withdrew day 5 n | Lost-to-follow-up day 30 n | Withdrew day 30 n |
|---|---|---|---|---|---|---|---|---|---|---|---|---|
| **Supportive Care** | | | | | | | | | | | | |
| A | 203 | 22 (10.8) | 22 (10.8) | 11 | 11 | | 22 (100) | 21 (95.5) | 0 | 0 | 1 | 0 |
| B | 2713 | 50 (1.8) | 33 (1.2) | 16 | 17 | | 33 (100) | 33 (100) | 0 | 0 | 0 | 0 |
| C | 2833 | 22 (0.8) | 21 (0.7) | 12 | 9 | | 21 (100) | 20 (95.2) | 0 | 0 | 1 | 0 |
| D | 3597 | 143 (4) | 118 (3.3) | 59 | 59 | | 118 (100) | 118 (100) | 0 | 0 | 0 | 0 |
| E | 4169 | 148 (3.6) | 103 (2.5) | 53 | 50 | | 102 (99) | 100 (97.1) | 0 | 1 | 2 | 0 |
| F | 1923 | 147 (7.6) | 106 (5.5) | 54 | 52 | | 106 (100) | 105 (99.1) | 0 | 0 | 1 | 0 |
| G | 8545 | 53 (0.6) | 18 (0.2) | 7 | 11 | | 18 (100) | 18 (100) | 0 | 0 | 0 | 0 |
| H | 363 | 4 (1.1) | 4 (1.1) | 2 | 2 | | 4 (100) | 4 (100) | 0 | 0 | 0 | 0 |
| I | 2405 | 85 (3.5) | 83 (3.5) | 42 | 41 | | 83 (100) | 83 (100) | 0 | 0 | 0 | 0 |
| J | 2352 | 57 (2.4) | 56 (2.4) | 28 | 28 | | 56 (100) | 56 (100) | 0 | 0 | 0 | 0 |
| K | 4912 | 155 (3.2) | 152 (3.1) | 77 | 75 | | 152 (100) | 152 (100) | 0 | 0 | 0 | 0 |
| L | 4495 | 156 (3.5) | 153 (3.4) | 76 | 77 | | 153 (100) | 152 (99.3) | 0 | 0 | 1 | 0 |
| M | 2957 | 49 (1.7) | 48 (1.6) | 24 | 24 | | 48 (100) | 47 (97.9) | 0 | 0 | 1 | 0 |
| N | 6609 | 136 (2.1) | 133 (2) | 65 | 68 | | 133 (100) | 131 (98.5) | 0 | 0 | 2 | 0 |
| O | 657 | 10 (1.5) | 9 (1.4) | 4 | 5 | | 9 (100) | 9 (100) | 0 | 0 | 0 | 0 |
| P | 608 | 7 (1.2) | 5 (0.8) | 2 | 3 | | 5 (100) | 5 (100) | 0 | 0 | 0 | 0 |
| **Total** | **49341** | **1244 (2.5)** | **1064 (2.2)** | **532** | **532** | | **1063 (99.9)** | **1054 (99.1)** | **0** | **1** | **9** | **0** |

[a] **Study sites:**

A- Nakuru Level 6 Hospital B- Thika Level 5 Hospital C- Jaramogi Oginga Odinga Teaching and Referral Hospital D- Naivasha County Referral Hospital E- Kiambu County Referral Hospital F- Machakos Level 5 Hospital G- Mama Lucy Kibaki Hospital H – Kerugoya County Referral Hospital I- Kisumu County Referral Hospital J- Kakamega County Teaching and Referral Hospital K- Busia County Referral Hospital L- Kitale County Referral Hospital M- Embu Level 5 Hospital N- Bungoma County Referral Hospital O- Kisii Teaching and Referral Hospital P-Kenyatta National Hospital

**Table S5 : Characteristics and outcomes of eligible participants who were not included in the trial.**

| Characteristics | Total<br>N = 482 |
|---|---|
| **History** | |
| Sex – *n (%)* | |
| *Male* | 271 (56.3) |
| *Female* | 210 (43.7) |
| Age (months) – *mean (SD)* | 17 |
| Re-admission – *n (%)* | 50 (10.4) |
| Discharged <1 month ago – *n (%)* | 14 (2.9) |
| Weight (kg) – *mean (SD)* | 8.0 (4.2) |
| Height (cm) – *mean (SD)* | 72.95 |
| WHZ – *mean (SD)* | 1.9 |
| MUAC (cm) (> 6months) – *mean (SD)* | 8.16 |
| Length of illness (days) – *mean (SD)* | 4.5 |
| Diarrhoea bloody - *Yes – n (%)* | 1 (0.1) |
| Vomiting – *Yes – n (%)* | 152 (31.5) |
| Freq. of vomiting in 24 hrs– *mean (SD)* | ***Not Captured*** |
| Difficulty feeding - *Yes – n (%)* | 166 (34.4) |
| Convulsions - *Yes – n (%)* | 22 (4.6) |
| **Immunization** | |
| Pneumo (doses) – *mean (SD)* | 0 ( |
| **Medication prior to admission** | |
| Antibiotics - *Yes – n (%)* | 405 (84.0) |
| **Examination** | |
| Temperature (degree Celsius) – *mean (SD)* | 37.3 |
| Respiratory rate (bmp) – *mean (SD)* | 16.0 |
| O2 saturation (%)– *mean (SD)* | 89 |

| | |
|---|---|
| Central Cyanosis - *Yes – n (%)* | 8 (1.7) |
| Indrawing - *Yes – n (%)* | 305 (63.3) |
| Wheezing - *Yes – n (%)* | 81 (16.8) |
| Grunting - *Yes – n (%)* | 142 (29.5) |
| Crackles - *Yes – n (%)* | 257 (53.3) |
| Capillary refill (/secs) – *n (%)* | ***Not Captured*** |
| Pallor Anaemia – *n (%)* | |
| *0* | 353 (73.2) |
| + | 60 (12.5) |
| +++ | 15 (3.1) |
| Can drink / breastfeed - *Yes – n (%)* | 360 (74.7) |
| **Diagnoses at admission** | |
| Malaria – *n (%)* | |
| *Severe* | 11 (2.3) |
| *Non-severe* | 2 (<1) |
| Diarrhoea – *n (%)* | |
| *Non-bloody* | 59 (6.1) |
| *Bloody* | 1 (<1) |
| *Not categorised* | 12 (1.2) |
| Dehydration – *n (%)* | |
| *Shock* | 5 (<1) |
| *Severe* | 14 (1.5) |
| *Some* | 34 (3.5) |
| HIV – *n (%)* | |
| *Positive* | 0 (0) |
| *Exposed / PMTCT+* | 7 |
| Malnutrition – *n (%)* | |
| *Mild* | 2 (<1) |
| *Moderate* | 14 (1.5) |
| *Severe* | 18 (1.9) |

| | |
|---|---|
| *Marasmus* | 2 (<1) |
| *Kwashiorkor* | 1 (<1) |
| Anaemia – *n (%)* | |
| *Severe* | 10 (1.0) |
| *Non-severe* | 16 (1.7) |
| Asthma – *n (%)* | |
| *Very severe* | 0 (0) |
| *Severe* | 7 (<1) |
| *Non severe* | 16 (1.7) |
| Suspected TB - *Yes – n (%)* | 5 (<1) |
| Sickle cell disease - *Yes – n (%)* | 3(0.31) |
| Meningitis - *Yes – n (%)* | 18 (1.9) |
| Rickets - *Yes – n (%)* | 26 (2.7) |
| **Outcomes** | |
| Mortality at 5 days *n (%)* | 33 (6.9) |
| Length of hospital stay *mean (SD)* | 5.59 (4.63) |

*Note: Percentages are column percentages*

**Table S6a: Per-protocol analysis of primary and secondary outcomes by antibiotic treatment arm – Factorial Analysis**

| | | | | Unadjusted | | | Adjusted** | | |
|---|---|---|---|---|---|---|---|---|---|
| **Outcome** | **Pen n (%)** | **Amox n (%)** | **Ceft n (%)** | **Amox vs Pen* RR (97.5% CI)** | **Ceft vs Pen* RR (97.5% CI)** | **Ceft vs Amox* RR (97.5% CI)** | **Amox vs Pen* RR (97.5% CI)** | **Ceft vs Pen* RR (97.5% CI)** | **Ceft vs Amox* RR (97.5% CI)** |
| **Mortality at 5 days** | 82 (5.90) | 80 (5.83) | 81 (5.74) | 0.99 (0.70, 1.39) p = 0.939 | 0.97 (0.69, 1.37) p = 0.858 | 0.98 (0.70, 1.39) p = 0.919 | 0.98 (0.70, 1.38) p = 0.899 | 0.98 (0.70, 1.38) p = 0.915 | 1.00 (0.71, 1.41) p = 0.984 |
| **Mortality at 30 days** | 114 (8.25) | 109 (8.00) | 119 (8.48) | 0.97 (0.73, 1.29) p = 0.809 | 1.03 (0.78, 1.36) p = 0.811 | 1.06 (0.80, 1.41) p = 0.631 | 0.96 (0.72, 1.27) p = 0.739 | 1.03 (0.78, 1.36) p = 0.807 | 1.08 (0.81, 1.42) p = 0.563 |
| **Experienced SAE** | 201 (14.47) | 203 (14.81) | 204 (14.48) | 1.02 (0.85, 1.23) p = 0.803 | 1.00 (0.84, 1.20) p = 0.995 | 0.98 (0.82, 1.17) p = 0.807 | 1.01 (0.85, 1.20) p = 0.917 | 1.00 (0.83, 1.19) p = 0.963 | 0.99 (0.83, 1.18) p = 0.880 |

| Outcome | Pen Median (IQR) | Amox Median (IQR) | Ceft Median (IQR) | Amox vs Pen* HR (97.5% CI) | Ceft vs Pen* HR (97.5% CI) | Ceft vs Amox* HR (97.5% CI) | Amox vs Pen* HR (97.5% CI) | Ceft vs Pen* HR (97.5% CI) | Ceft vs Amox* HR (97.5% CI) |
|---|---|---|---|---|---|---|---|---|---|
| **Length of hospital stay** | 4 (3, 6) | 4 (3, 6) | 4 (3, 6) | 1.00 (0.91, 1.08) p = 0.913 | 1.02 (0.94, 1.11) p = 0.568 | 1.03 (0.94, 1.12) p = 0.497 | 0.99 (0.91, 1.08) p = 0.851 | 1.01 (0.92, 1.10) p = 0.851 | 1.01 (0.93, 1.10) p = 0.723 |
| **Duration to tolerate full fluid requirements by mouth** | 0 (0, 1) | 0 (0, 1) | 0 (0, 1) | 1.00 (0.87, 1.16) p = 0.955 | 1.01 (0.87, 1.16) p = 0.922 | 1.00 (0.86, 1.16) p = 0.968 | 0.98 (0.85, 1.14) p = 0.782 | 0.98 (0.85, 1.14) p = 0.799 | 1.00 (0.86, 1.16) p = 0.983 |
| **Time to death or discharge** | 4 (3, 6) | 4 (3, 6) | 4 (3, 6) | 1.00 (0.91, 1.09) p = 0.913 | 1.02 (0.94, 1.11) p = 0.568 | 1.03 (0.94, 1.12) p = 0.497 | 0.99 (0.91, 1.08) p = 0.868 | 1.01 (0.92, 1.10) p = 0.851 | 1.01 (0.93, 1.10) p = 0.723 |

** Comparison group*

*** Adjusted for age, site and feeding arm; note that this limits the sample to only those randomised to both interventions*

**Table S6b: Per-protocol analysis of primary and secondary outcomes by supportive care treatment arm – Factorial Analysis**

| | | | Unadjusted | Adjusted** |
|---|---|---|---|---|
| **Outcome** | **NG**<br>**n (%)** | **IV**<br>**n (%)** | **RR* (95% CI)** | **RR* (95% CI)** |
| **Mortality at 5 days** | 29 (5.70) | 33 (6.42) | 1.13 (0.69, 1.83)<br>p = 0.628 | 1.09 (0.68, 1.74)<br>p = 0.715 |
| **Mortality at 30 days** | 34 (6.75) | 40 (7.83) | 1.16 (0.75, 1.80)<br>p = 0.508 | 1.16 (0.76, 1.77)<br>p = 0.487 |
| **Experienced SAE** | 55 (10.81) | 67 (13.06) | 1.21 (0.86, 1.69)<br>p = 0.267 | 1.17 (0.85, 1.63)<br>p = 0.340 |
| | **NG**<br>**Median (IQR)** | **IV**<br>**Median (IQR)** | **HR (95% CI)** | **HR (95% CI)** |
| **Length of hospital stay** | 4 (3, 5) | 4 (3, 6) | 0.95 (0.84, 1.07)<br>p = 0.376 | 0.96 (0.85, 1.09)<br>p = 0.547 |
| **Duration to tolerate full fluid requirements by mouth** | 1 (1, 2) | 1 (1, 2) | 0.93 (0.82, 1.06)<br>p = 0.308 | 0.93 (0.82, 1.06)<br>p = 0.275 |

| | | | | |
|---|---|---|---|---|
| **Time to death or discharge** | 4 (3, 5) | 4 (3, 6) | 0.95 (0.84, 1.07)<br><br>p = 0.376 | 0.96 (0.85, 1.09)<br><br>p = 0.546 |

**NG is comparison group*

*** Adjusted for age, site and antibiotic arm*

**Figure S4a Between Hospital Comparison for Day 5 Mortality- Benzylpenicillin plus gentamicin vs Ceftriaxone**

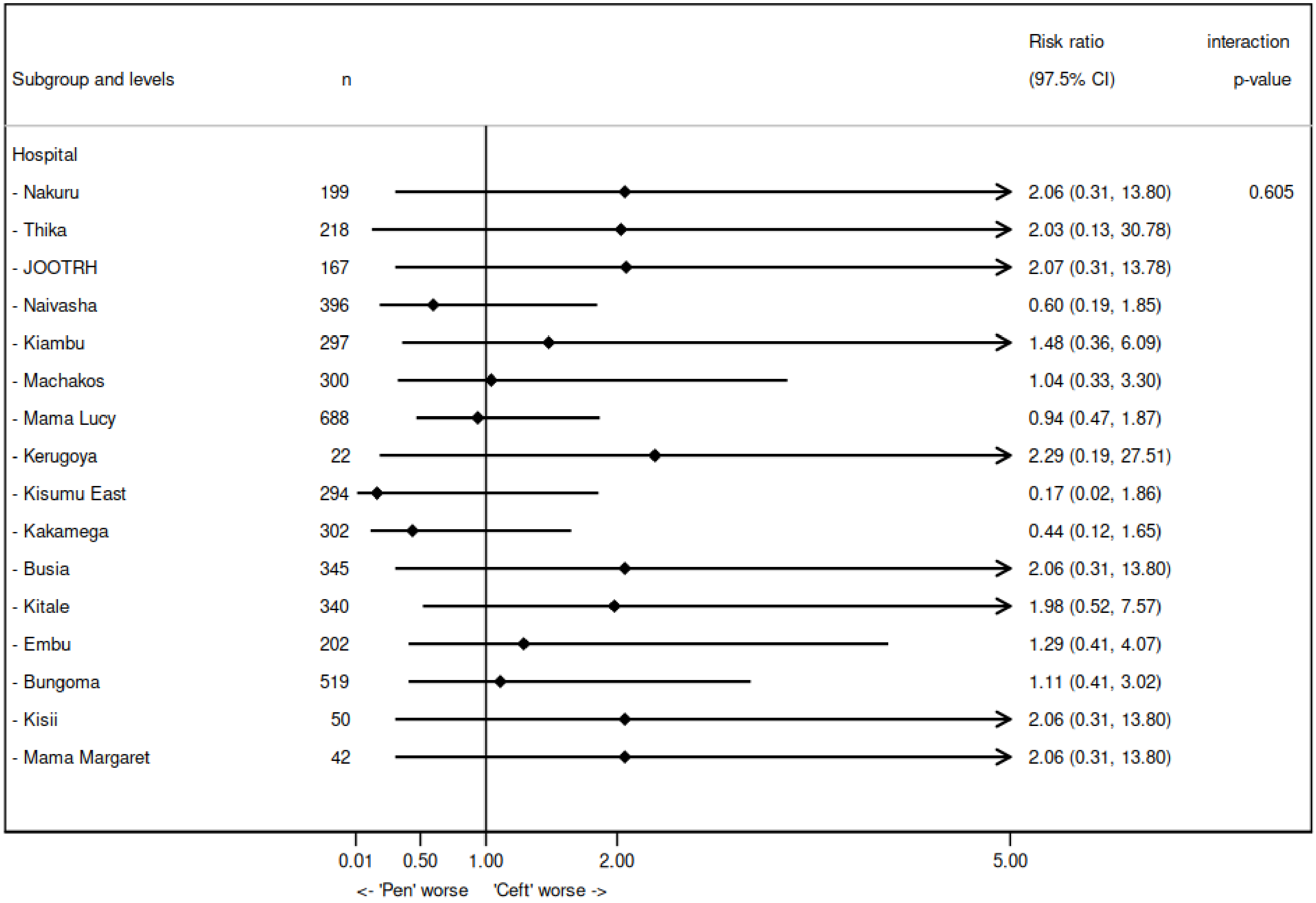

**Figure S4b Between Hospital Comparison for Day 5 Mortality- Benzylpenicillin plus gentamicin vs Amoxicillin-clavulanic acid**

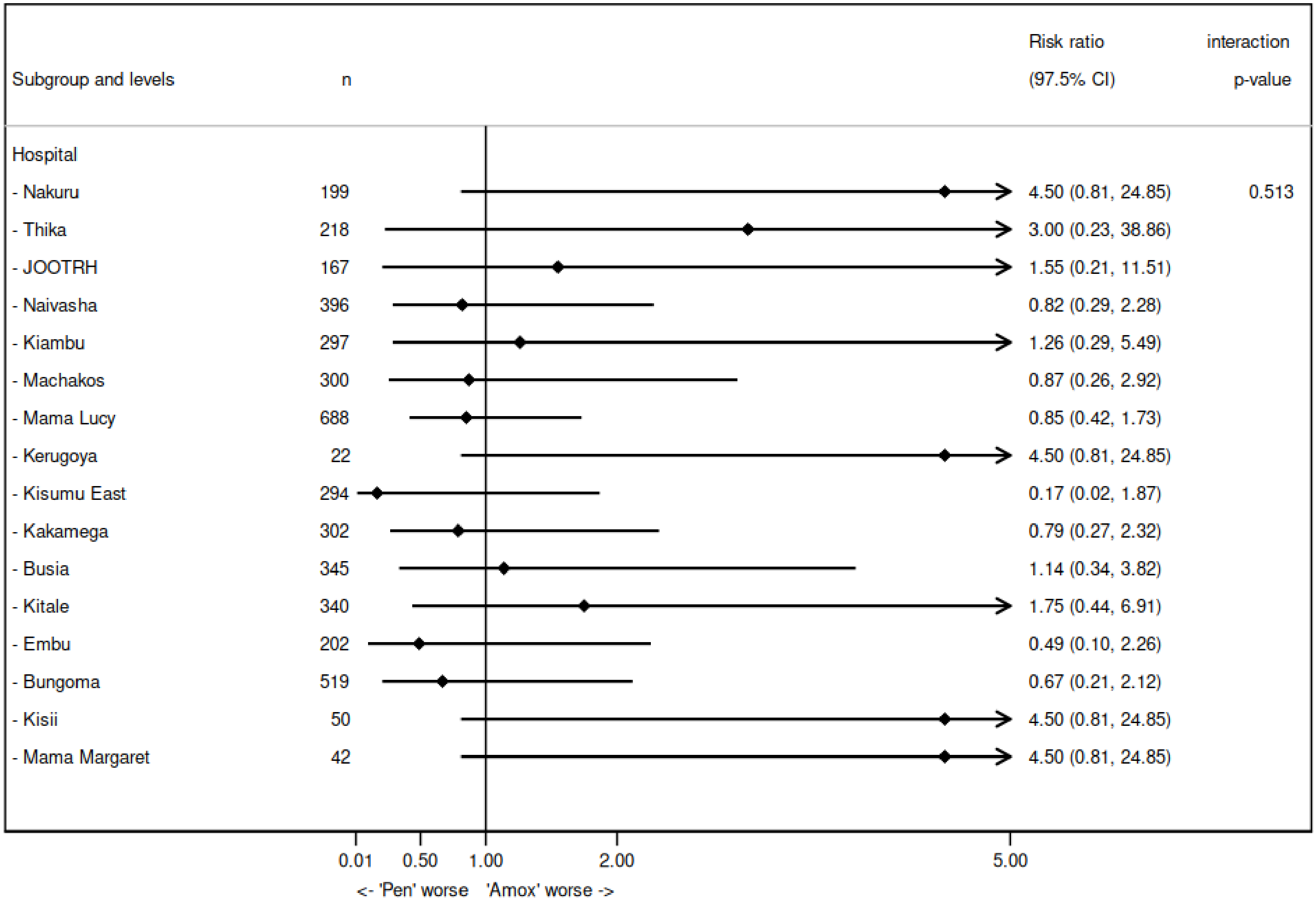

**Figure S4c Between Hospital Comparison for Day 5 Mortality- Ceftriaxone vs Amoxicillin-clavulanic acid**

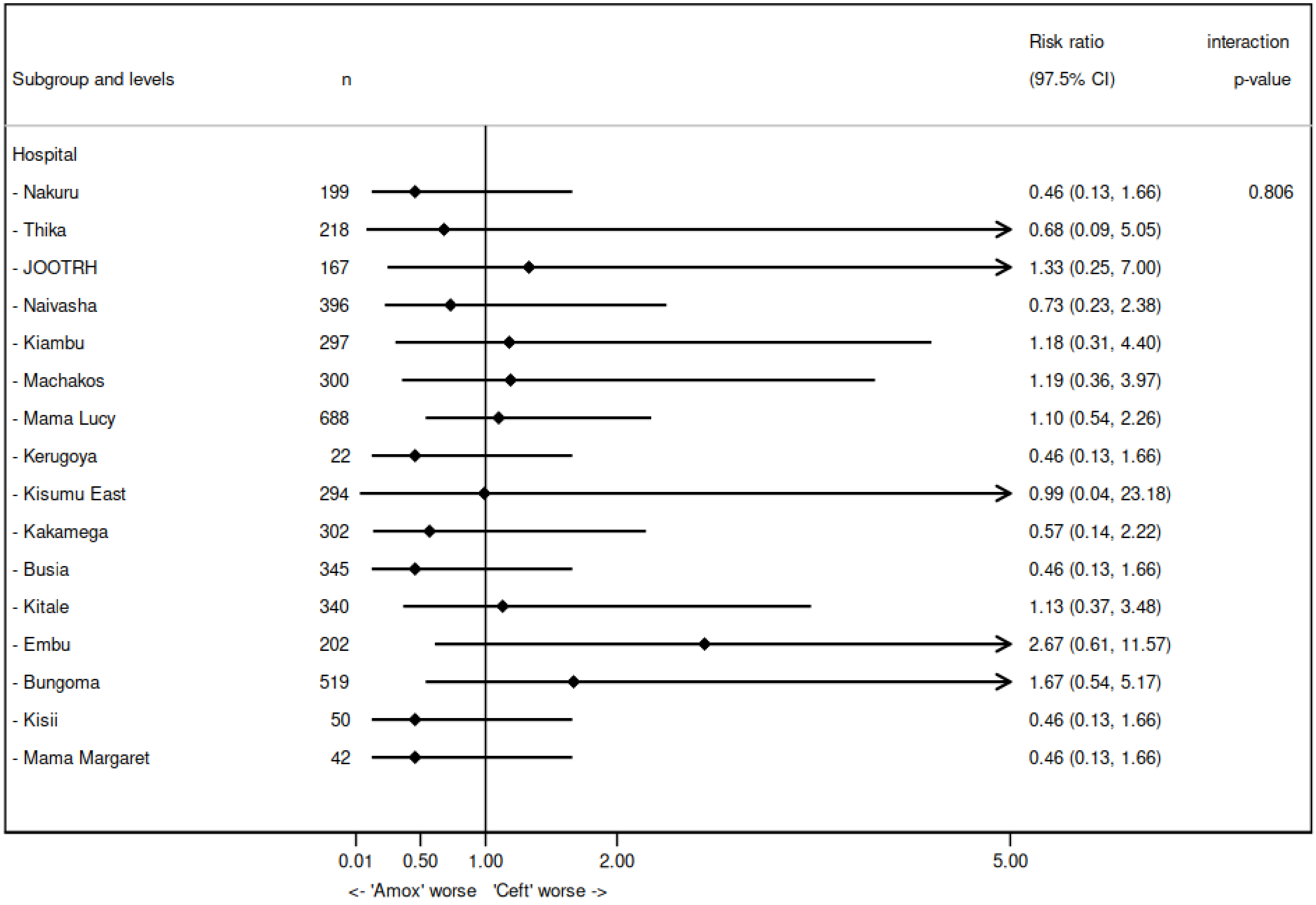

**Figure S4d Between Hospital Comparison for Day 5 Mortality- Intravenous fluids vs Nasogastric tube feeds**

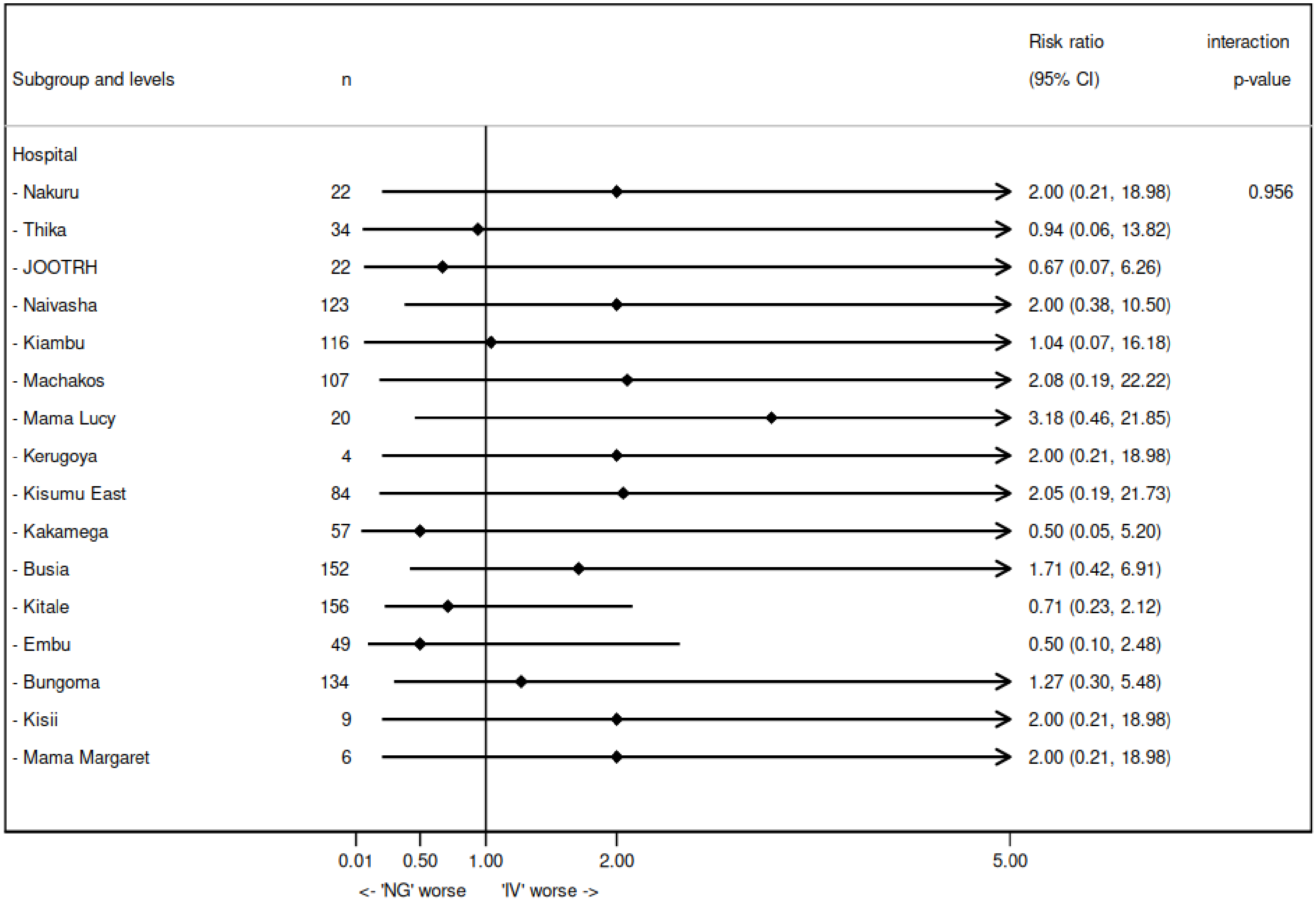

**Table S7 Complier-average causal effect estimates for the effects of the antibiotic and supportive factors on the primary outcome**

| | Unadjusted | | | | Adjusted** | | | |
|---|---|---|---|---|---|---|---|---|
| **Outcome** | **AMX-CLV vs PEN-GENT * RR (97.5% CI)** | **CEF vs PEN-GENT * RR (97.5% CI)** | **CEF vs AMX-CLV * RR (97.5% CI)** | **IVF vs NGT*** | **AMX-CLV vs PEN-GENT * RR (97.5% CI)** | **CEF vs PEN-GENT * RR (97.5% CI)** | **CEF vs AMX-CLV * RR (97.5% CI)** | **IVF vs NGT*** |
| **Mortality at 5 days** | 0.67 (0.40, 1.12) p = 0.129 | 0.98 (0.44, 2.13) p = 0.953 | 1.45 (0.62, 3.40) p = 0.387 | 1.18 (0.49, 2.86) p = 0.712 | 0.69 (0.41, 1.15) p = 0.153 | 1.07 (0.48, 2.36) p = 0.875 | 1.54 (0.66, 3.63) p = 0.318 | 1.17 (0.48, 2.87) p = 0.727 |

** Comparison group*

*** Adjusted for age and feeding arm; note that this limits the sample to only those randomised to both interventions. Further adjustment for site resulted in model non-convergence*
